# Commuting-driven competition between transmission chains shapes seasonal influenza virus epidemics in the United States

**DOI:** 10.1101/2024.08.09.24311720

**Authors:** Simon P.J. de Jong, Andrew Conlan, Alvin X. Han, Colin A. Russell

## Abstract

Despite intensive study, much remains unknown about the dynamics of seasonal influenza virus epidemic establishment and spread in the United States (US) each season. By reconstructing transmission lineages from seasonal influenza virus genomes collected in the US from 2014 to 2023, we show that most epidemics consisted of multiple distinct transmission lineages. Spread of these lineages exhibited strong spatiotemporal hierarchies and lineage size was correlated with timing of lineage establishment in the US. Mechanistic epidemic simulations suggest that mobility-driven competition between lineages determined the extent of individual lineages’ geographical spread. Based on phylogeographic analyses and epidemic simulations, lineage-specific movement patterns were dominated by human commuting behavior. These results suggest that given the locations of early-season epidemic sparks, the topology of inter-state human mobility yields repeatable patterns of which influenza viruses will circulate where, but the importance of short-term processes limits predictability of regional and national epidemics.

**Teaser:** Epidemics consist of multiple sub-epidemics that compete for susceptible hosts and spread due to the movement of commuters.

## Introduction

In the United States, seasonal influenza epidemics recur every year as the result of a complex hierarchy of transmission processes. Intercontinental viral migration, driven by global metapopulation dynamics, drives the initial early-season seeding of epidemics in the US (*1–3*). Following these initial epidemic sparks, inter-state patterns of human mobility disseminate viruses across the country (*4–9*), resulting in an interconnected network of local epidemics (*10*, *11*). These epidemics vary substantially from year to year in their timing, size, and composition (*12–14*). Gaining a predictive understanding of the variables that shape the composition, timing and magnitude of these epidemics is a key public health target (*15*). Substantial efforts have been put into forecasting the timing of epidemic onset and epidemic peaks to aid public health planning (*14*, *16–21*).

Knowledge of the underlying transmission processes that give rise to epidemic establishment and subsequent spread is essential for a predictive understanding of epidemic characteristics (*22*, *23*). For example, does peak-period epidemic activity arise from the gradual expansion of early-season transmission chains, or are epidemics the result of transmission chains that rapidly expanded when conditions became favorable for large-scale transmission? Similarly, do epidemics tend to comprise a single epidemic wave that sweeps across country, or rather do they consist of many co-circulating transmission lineages that jointly shape epidemics (*24*, *25*)? Further questions remain regarding the underlying mobility drivers of viral spread, such as the roles of air travel and commuting in disseminating viruses country-wide (*4–6*, *26*). The US forms a particularly compelling setting to explore fundamental questions about the determinants of influenza virus spread due to its geographical expanse, climatic variability and complex mobility networks.

Most previous studies into seasonal influenza epidemic dynamics in the US have relied primarily on virological and syndromic surveillance data, such as pneumonia and influenza (P&I) mortality data or influenza-like illness (ILI) data (*4–6*). However, such data cannot effectively distinguish between distinct chains of transmission, potentially limiting the precision and specificity with which the underlying dynamics of epidemic establishment and viral migration can be reconstructed (*4*, *23*). Hence, we turned to genomic data, collected during routine surveillance in the United States. By decomposing epidemics into contributions of individual transmission lineages and reconstructing their individual spread, we aimed to gain more fine-grained insight into the processes of epidemic establishment and spread.

## Results

### Influenza virus epidemics consist of many distinct co-circulating transmission lineages

First, we characterized the transmission lineage structure of US seasonal influenza epidemics. We investigated whether epidemics tend to comprise many distinct co-circulating transmission lineages that independently emerged in different states, or rather consist of a single dominant transmission lineage that propagates across the country. We analyzed 30,508 whole-genome seasonal influenza virus sequences from the 48 contiguous states and the District of Columbia, collected during routine surveillance in the United States from 2014 to 2023, the most recent period for which substantial whole-genome sequences were available. In this period, all four influenza A subtypes/influenza B lineages (henceforth, subtypes) caused epidemic activity, but patterns of subtype dominance differed substantially from season to season (Fig. S1). To classify the viruses circulating in each season into transmission lineages, we phylogenetically grouped the viruses into clusters of viruses that exhibit a comb-like branching structure, suggestive of exponential spread (*27*). Given the exponential nature of influenza virus epidemics, we posit that groups of viruses with such a rapidly expanding branching structure plausibly represent groups of viruses that expanded from a single ancestral virus in the United States (Fig. 1A, S2-5).

**Fig. 1:**
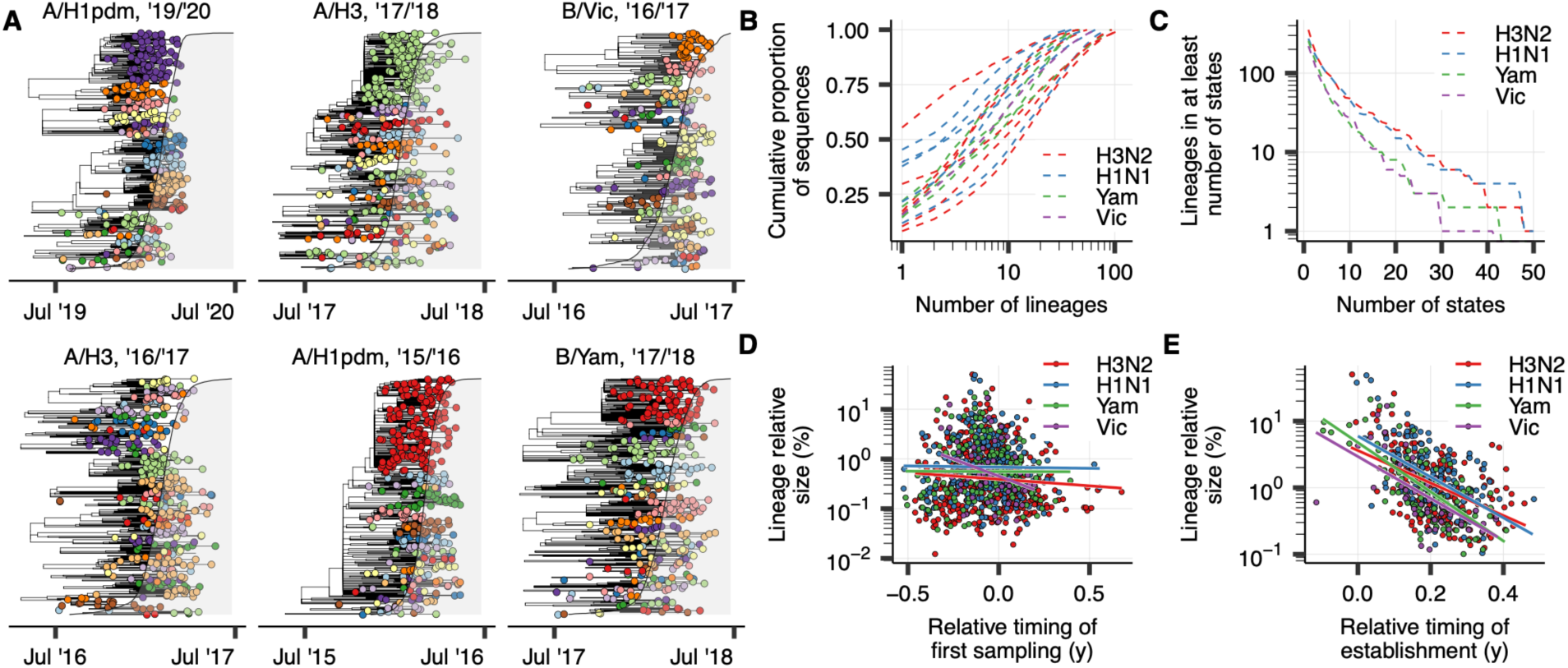
Lineage structure of US seasonal influenza epidemics. (A) Phylogenies of six representative subtype-season pairs, with tips colored by identified transmission lineage. The shaded grey area corresponds to the cumulative proportion of nation-wide positive tests in public health laboratories of the corresponding subtype at each point in time. (B) The size distribution of lineages by season and subtype. Each line represents the cumulative proportion of sequences that is accounted for by a number of lineages on the *x*-axis. Each line corresponds to an individual season, for an individual subtype. (C) The number of lineages across that accounts for >5% of sequences in a season-subtype in at least the number of states on the *x*-axis, by subtype. (D) Relationship between the first collection date of virus in a lineage and the lineage’s country-wide size normalized by state. Lineage sampling dates were computed relative to the timing of nation-wide epidemic onset, which was defined as the first week in which >5% of the season’s cumulative positive tests had been collected. (E) Relationship between the timing of establishment of substantial circulation of a lineage and its country-wide size normalized by state. Lineage establishment timing was computed relative to nation-wide epidemic onset analogous to (D).

Using this procedure, we clustered 81.2% of sequences into 3,842 lineages of at least two viruses. In most seasons, a relatively small number of transmission lineages accounted for the bulk of sequenced viruses (Fig. 1B), with the median minimum number of lineages that together accounted for at least 50% of sequences amounting to 5 lineages (range 1-13) across all seasons for subtypes that accounted for >10% of positive tests in the respective season. The degree of lineage diversity differed substantially across seasons (Fig. 1B). For example, in the 2015/2016 A/H1N1pdm09 epidemic, a single transmission lineage accounted for >50% of sequenced viruses, normalized across states. In contrast, in the 2016/2017 A/H3N2 season, the largest lineage accounted for only 6.4% of sequenced viruses. Lineage structure was evident for both circulating influenza A virus subtypes and both influenza B virus lineages, though transmission lineage clustering results are likely more error-prone for influenza B viruses given their lower evolutionary rate (*28*), particularly in seasons that saw relatively little circulation and were less densely sampled.

Consistent with the lineage size distribution, most transmission lineages were confined to a relatively small number of states, with a small proportion of lineages spreading widely across the country (Fig. 1C): among the 1,104 identified transmission lineages that accounted for at least 5% of sequences in a season in at least one state, 144 (13.0%) lineages did so in at least 10 states, and 27 (2.4%) did so in at least 25. Patterns of lineage diversity at the state level mirrored those at the national level, with some seasons seeing very high within-state lineage diversity (e.g. 2018/2019 A/H1N1pdm09, median state-wise Shannon entropy of lineage composition = 0.76, inter-quartile range 0.68-0.82), whereas in other seasons a few lineages dominated state-level epidemics (e.g. 2018/2019 A/H3N2, median Shannon entropy = 0.44, inter-quartile range = 0.31-0.58). These results indicate that in most seasons, seasonal influenza epidemics are the result of the co-circulation of multiple independent chains of transmission, consistent with previous studies into individual seasons, both at the national and state level (*24*, *25*).

### Lineage size correlates with timing of establishment but not emergence

Next, we investigated the factors that influence the extent to which any individual transmission lineage will spread country-wide. We hypothesized that onset timing would explain the substantial variation in lineage size, where the first lineages to emerge in any season, for any subtype, would be larger. Here, we defined lineage size as the proportion of sequences that a lineage accounts for in a season for a subtype across all states, where each state has an equal weight. However, across all subtypes and seasons, we found that a relationship between time of first sampling of a lineage and (log) lineage size was weak (Spearman *ρ* = -0.07, *P* = 0.024) (Fig. 1D). We observed the proliferation of some transmission lineages that were first sampled a substantial amount of time prior to onset of nation-wide epidemic activity, but many of the most successful lineages emerged and were first sampled relatively close in time to the ramp-up of national epidemic activity (Fig. 1D).

For example, by the time of first sampling of the largest lineage in the highly severe (*29*) 2017/2018 season (Fig. 1A, topmost lineage), >10% of all the season’s sequences had already been collected. Despite its relatively late emergence, the lineage accounted for >40% of sequences during peak epidemic periods following rapid expansion. These rapidly expanding lineages could in some cases descend from unsampled viruses that had circulated prior locally at low levels, but the fact that a single ancestral virus could rapidly sweep to national dominance despite emerging at a time when many other transmission lineages already circulated suggests that early-season transmission processes are highly heterogeneous. Furthermore, the fact that the lineages dominating during peak epidemic periods often rapidly expanded around the time of epidemic onset, outcompeting co-circulating low-level transmission lineages, indicates that in many seasons very short-term epidemiological processes are crucial determinants of seasonal influenza epidemic dynamics.

The weak correlation between timing of first lineage sampling and lineage size suggests that early-season transmission chains frequently go extinct before the onset of substantial epidemic activity. However, we hypothesized that if a lineage did cause substantial epidemic activity early on, it would be well-positioned to be successful country-wide. Correspondingly, we found that the timing of establishment of substantial epidemic activity of a transmission lineage correlated strongly with nation-wide lineage size (Spearman *ρ* = -0.53, *P* < 0.001) (Fig. 1E). Here, we defined the timing of lineage establishment as the first week in which the lineage accounted for substantial epidemic activity (i.e. at least 5% of total estimated incidence in the season; see Materials and Methods) in at least one state. The fact that the lineages that first established substantial epidemic activity somewhere in the US were more likely to be successful country-wide suggests that the states with the earliest epidemic onset have outsized contributions to nationwide epidemic lineage composition.

### Transmission lineages are highly spatially structured

To investigate the extent to which transmission lineages are spatially structured, we computed the Bray-Curtis similarity index of epidemic transmission lineage compositions for all pairs of states. Here, states that more frequently sampled viruses belonging to the same transmission lineages have a higher similarity index. Aiming to identify communities of states that are more closely linked to one another than to other states, we performed hierarchical clustering on the similarity matrices. Qualitatively, this clustering recapitulated the geography of the United States, with relatively higher similarity for states within the same census region (Fig. 2A). Projecting the similarities among states onto a two-dimensional surface further recapitulated this spatial structure; for example, states belonging to the Northeast and Southeast appeared to form distinct clusters (Fig. 2B). However, the continuous distribution of states on the plane suggests states cannot consistently be classified into distinct communities, suggestive of substantial inter-regional mixing.

**Fig. 2:**
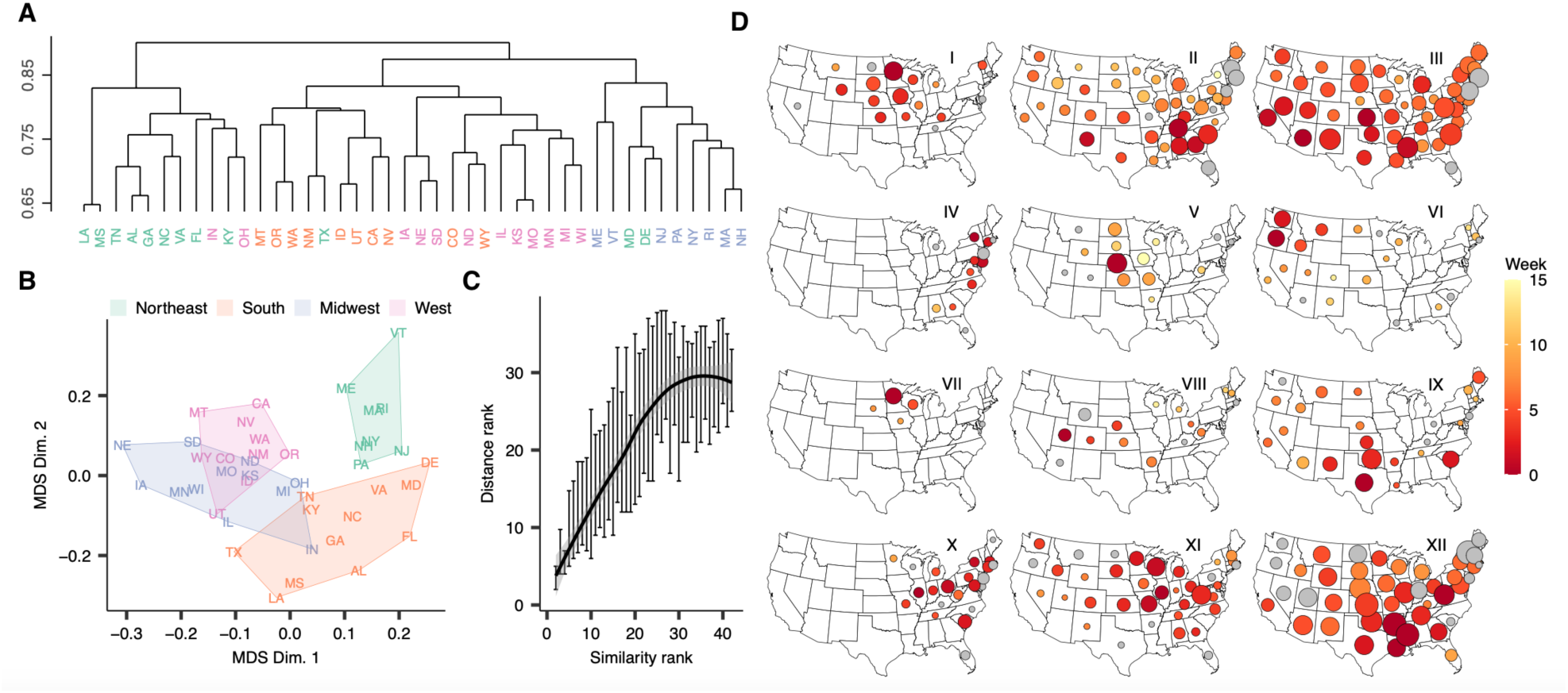
Spatial structure of US seasonal influenza epidemics. **(A)** Complete-linkage hierarchical clustering of pairwise state-to-state transmission lineage composition Bray-Curtis similarities across all subtypes, colored by census region. **(B)** Multi-dimensional scaling plot of the pairwise lineage composition Bray-Curtis similarity among states, colored by census region. **(C)** Relationship between pairwise transmission lineage compositional similarity and pairwise centroid distance rank. Vertical lines show 50% CI for each value of rank similarity, line corresponds to LOESS fit to medians. **(D)** Examples of lineage spatiotemporal spread. In each map, circle size and color correspond to the relative size and establishment timing of the corresponding lineage in each state, respectively. Grey fill corresponds to unknown lineage establishment timing.

Consistent with the states’ clustering by geography, we found that epidemics in states in closer geographic proximity more frequently comprised the same transmission lineages (Mantel test, *P* < 0.001) (Fig. 2C). The highest similarity indices were found for neighboring states (highest: MS-LA, MO-KS, GA-AL, NH-MA, UT-ID), with a neighboring state being the state with the highest similarity in 81% (34/42) of states included in the analysis. Stratifying by season and subtype, this correlation between distance and similarity (Mantel test, *P* < 0.01) was present in almost all (14/15) subtype-season pairs that accounted for >10% of nation-wide positive tests in their respective season. Together, these results show that at the transmission lineage level, US seasonal influenza epidemics are highly spatially structured.

To further investigate the spatiotemporal dynamics of viral spread, we reconstructed the spread of individual transmission lineages. We mapped the sampling dates of the viruses in each lineage to epidemiological data to quantify, in each state, 1) the relative size of each lineage, defined as the proportion of all viruses of that subtype that was accounted for by the lineage in that season; and 2) the week of lineage establishment, defined as the first week the lineage accounted for substantial levels of circulation in that state (i.e. at least 5% of total estimated incidence in the season; see Materials and Methods). We visualized lineage spread by projecting the timing of establishment and size of the lineage in each state on a map. These visualizations revealed a striking landscape of seasonal influenza spatial spread at the transmission lineage level, with examples for a geographically and temporally representative set of lineages shown in Fig. 2D. We identified instances of lineage emergence from all regions of the US, each with distinct signatures of spread. Across seasons and subtypes, many lineages exhibited a radial spatiotemporal progression and were highly regional (e.g. I, V, VI, VII, Fig. 2D); other lineages saw rapid cross-country spread (e.g. II, III, XII, Fig. 2D).

### Consistent source-sink dynamics are absent across seasons and subtypes

The above results suggest that lineages can potentially emerge in any region of the United States. We sought to more rigorously investigate seasonal influenza virus source-sink dynamics at the transmission lineage level for all seasons and subtypes by identifying each lineage’s most likely origin state. We performed discrete trait phylogeographic reconstructions for each lineage individually in BEAST (*30*), identifying the Health & Human Services (HHS) region that represented the most likely region of initial expansion for each of the 262 transmission lineages that accounted for >0.5% of sampled viruses, normalized by state, in their respective season. To ensure our results were robust to differences in sampling across regions, we performed these analyses with two distinct subsampling strategies: first, one where the number of sequences included for each HHS region depended on its population size; and second, one with a constant number of sequences across HHS regions. We found substantial season-to-season variation for the most probable origin regions for successful lineages. Of the seven transmission lineages that accounted for >10% of sequences in a single season, normalized across states, three likely first established in the South (HHS regions 4 & 6; e.g. lineages II & XII, Fig. 2D), one likely emerged in the West (HHS region 9; lineage III, Fig. 2D), one in the Midwest (HHS regions 7 & 5; lineage XI, Fig. 2D), one in the Northeast (HHS region 1), and one could not consistently be attributed to a single region across both sampling strategies.

With the aim of identifying regional variation in source-sink dynamics, we computed state-specific origin profiles that quantify the role of each HHS region as the region of initial lineage expansion of sampled viruses in each state. These profiles differed substantially across states (Fig. S6, S7). For example, averaged across both subsampling strategies, the proportion of sequences reconstructed to coalesce to epidemic expansions in HHS region 4, encompassing most of the Southeast, ranged from 39.3% in South Carolina (HHS region 4) and 27.4% in Arkansas (HHS region 6) to 13.0% in Arizona (HHS region 8). Similarly, lineages expanding in HHS region 10, encompassing the Pacific Northwest, accounted for 15.3% of sampled viruses in Idaho (HHS region 10), 10.7% in North Dakota (HHS region 8), and only 2.7% in Arkansas (HHS region 6).

States in closer geographic proximity saw more similar origin profiles, even if they corresponded to different HHS regions (Mantel test, *P* < 0.001). Across all states, a relatively limited proportion of viruses corresponded to lineages that originated from the state’s own HHS region (median 17.3%, range 11.4%-29.8% for uniform subsampling strategy), suggesting a high degree of viral mixing at the national level. Importantly, origin profiles were strongly correlated across the two subsampling strategies (Spearman *ρ* = 0.81, *P* < 0.001). Together, these results suggest that influenza virus source-sink dynamics are highly heterogeneous, without consistent source regions of successful lineages, but are spatially structured.

### Mechanistic simulations suggest commuting flows drive viral spread

Our analyses established a strong correlation between timing of lineage establishment and lineage size (Fig. 1E). However, this correlation does not account for a substantial portion of the observed variation in transmission lineage size. We hypothesized that differences in mobility flows, coupled to inter-lineage competition, could explain why some lineages spread widely following local establishment, whereas other lineages remain highly spatially constrained. Importantly, the reconstructed spread of individual lineages provided a vital ground truth which we could leverage to resolve long-standing questions regarding the underlying mobility determinants of influenza virus spread.

The lineage competition hypothesis appears to explain lineage dynamics in the 2018/2019 A/H3N2 season. The beginning of this season was dominated by A/H1N1pdm09 viruses, but it also saw the rapid expansion of viruses of the A/H3N2 subtype that were associated with decreased vaccine effectiveness (*31*). Phylogenetic analyses, integrated with epidemiological data, indicate that A/H3N2 circulation in this season was dominated by two lineages, appearing to emerge from Georgia (GA, lineage 1) and Nebraska (NE, lineage 2), respectively (Fig. 3, left), each establishing swiftly as evidenced by a comb-like, rapid branching structure. Visualizations of the lineages’ distributions across states suggest that spread of the Nebraskan lineage was regional and radial, causing substantial epidemic activity mainly in the immediately surrounding states in a clear spatiotemporal hierarchy (Fig. 3, left). Conversely, the lineage from Georgia quickly spread to almost all states with a less prominent temporal hierarchy, although it appeared to arrive in neighboring states first. We hypothesized that competition from the Georgian lineage explained why spread of the Nebraskan lineage remained so regional. In turn, this would explain why the Georgian lineage failed to spread substantially in Nebraska and immediately surrounding states.

**Fig. 3:**
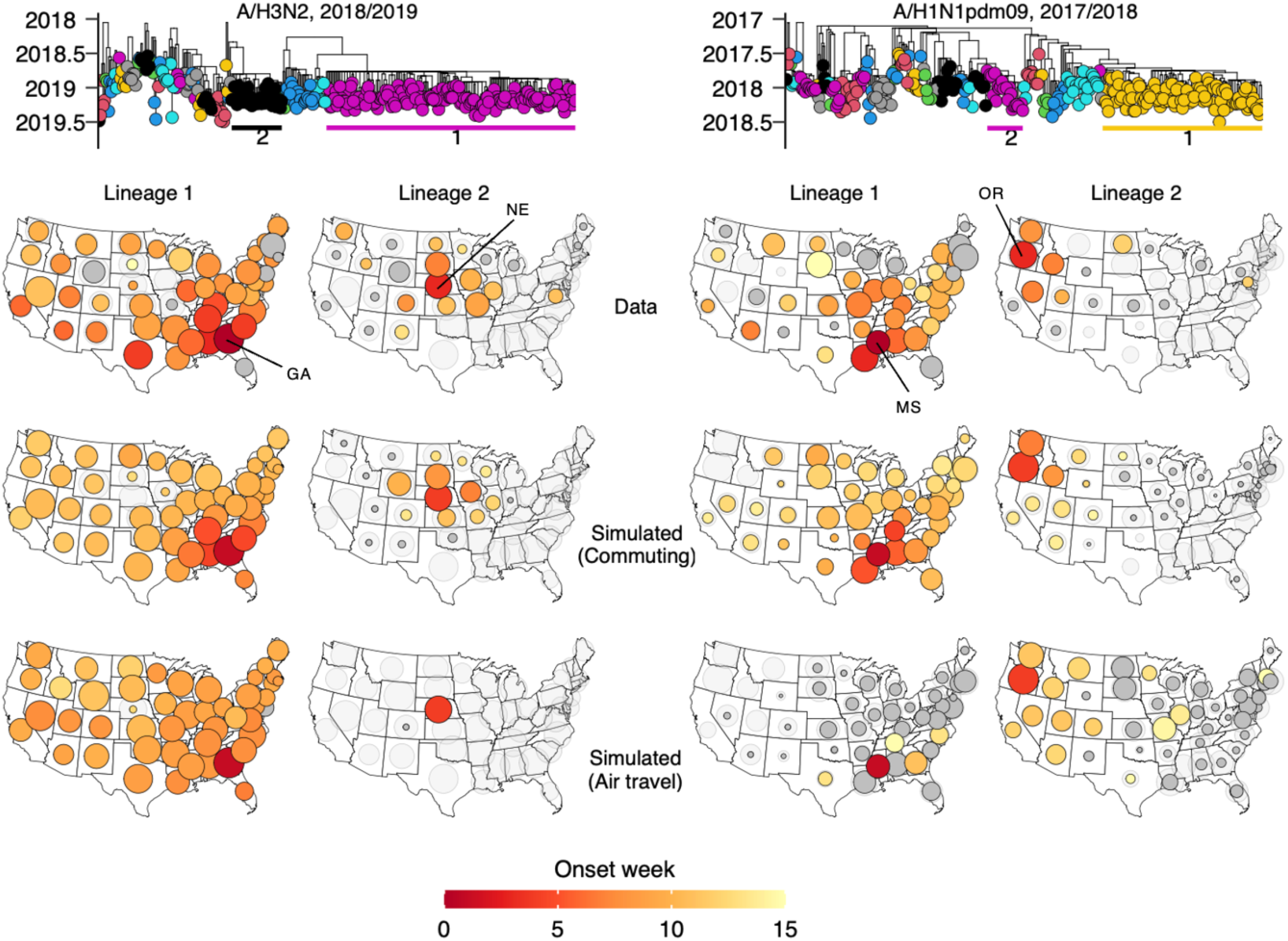
Mobility drivers of influenza virus spread. Phylogenies represents the 2018/2019 A/H3N2 season (left) and 2017/2018 A/H1N1pdm09 season (right), with the two largest lineages labeled in each. For both seasons, the maps in the top row visualize the reconstructed spread of the labeled lineages, with size and color corresponding to lineage size and establishment timing, in each state, respectively. Middle maps show simulated spread of the two lineages for each of the two seasons, using commuting data, when initialized in their origin state in their likely origin week. Bottom maps are analogous to the middle maps but using air travel data instead of commuting data. Light grey circles represent the total proportion of sequences in that state that are accounted for by the lineages that were simulated, to account for the fact that simulations only incorporated a subset of all lineages; circles for the simulated lineages have their size scaled such that the sum of simulated lineages’ sizes for each state is proportional to the proportion of sequences accounted for by the simulated lineages in that state (i.e., the light grey area). Dark grey fill corresponds to absence of an establishment week (for top row, potentially due to missing data), or establishment after the 15^th^ week.

To test the hypothesis that competitive interactions between lineages, coupled to mobility, drive lineage spread, we explored if we could reproduce the spread of these two lineages in mechanistic epidemic simulations. To do so, we used a metapopulation model that models viral spread between states in a susceptible-infected-recovered (SIR) epidemic framework. The model simulates the spread of multiple co-circulating lineages that compete for disease-susceptible individuals with perfect cross-immunity between transmission lineages. We initialized the simulations in the lineages’ respective establishment weeks, in their respective onset states (Georgia and Nebraska) and simulated forward in time to model the spread of the two lineages. By visualizing which of the two lineages would predominate in each state in the simulations, we could ascertain if we could reproduce their observed spread. To ascertain the predominant mobility drivers of viral spread, we parameterized rates of state-to-state mobility using either commuting flows, extracted from the US Census Bureau, or air travel data, extracted from the US Department of Transportation.

When using rates of commuting to parameterize rates of inter-state travel, we could reproduce the observed spread of the two lineages with striking accuracy: the simulations recapitulated the radial spread from Nebraska, and the relative success of the lineage emerging from Georgia (Figure 3, left). The simple model of inter-lineage competition driven by commuting also explains why the lineage from Georgia failed to cause epidemic activity in the Nebraska and the immediately surrounding states. On the other hand, the correspondence to observed spread was very poor when using air travel flows, with an absence of substantial spread from Nebraska. Together, these mechanistic simulations suggest that commuting flows were the primary correlate of viral spread. These results also support the notion that competitive interactions between lineages, mediated by mobility flows, shape the distribution of lineages across states.

### Spatial segregation and limited competition allow lineages from small states to spread widely

The 2018/2019 A/H3N2 season lends genome-informed credence to the conjectured gravity-like spread of seasonal influenza viruses (*5*), with a lineage originating from a populous, highly connected state (in this case, Georgia, population ∼11 million) spreading quickly through strong long-range connections, while spread from a smaller state (Nebraska, population ∼2 million) was slower and more local. Georgia’s high degree of connectivity and earlier onset allowed lineage 1 to spread to other states more rapidly than lineage 2, with its day of arrival in another state on average 42 days (IQR 27-53) earlier than lineage 2’s in metapopulation simulations. Nevertheless, the substantial spread of the Nebraskan lineage shows that spatial segregation between lineages can allow a lineage emerging from a small state to proliferate, even if it co-circulates with a lineage emerging from a more populous state, as long as it sees sufficiently early establishment and is spatially segregated from the lineage emerging from the larger state.

Our results suggests that by facilitating spread from less populous states, the short-range spatial coupling reflected in commuting flows is a key determinant of seasonal influenza virus spread. This notion is further supported in the 2017/2018 A/H1N1pdm09 season, in which the two largest lineages appeared to emerge in Mississippi (MS) and Oregon (OR), respectively (Fig. 3, right). When using commuting flows, the relative degree of spread of the two lineages could be reproduced. Despite its relatively small population, Mississippi’s high connectivity through commuting flows allowed lineage 1 to rapidly spread beyond local constraints. In contrast, due to Oregon’s relatively limited connectivity and the later establishment of lineage 2, competition from lineage 1 likely constrained the spread of those viruses to the Western United States. When using only air travel to parameterize inter-state mobility, the simulations strongly overestimated the degree of spread from Oregon relative to Mississippi, with too slow spread from Mississippi, compared to the ground truth (Fig. 3, right).

Using counterfactual simulations, we explored how mobility interacts with establishment timing to competitively shape the spread of individual lineages. Under the baseline simulations for the 2018/2019 A/H3N2 season, lineage 2 accounted for >10% of circulation among the two lineages in 11 states. Simulations indicate that had lineage 2 established in Nebraska four weeks later (with lineage 1’s establishment timing unchanged), lineage 2 would have accounted for >10% of circulation in only 4 states, constrained by competition from lineage 1. Conversely, if it had established four weeks earlier, lineage 2 would have been accounted for >10% of circulation in 37 states, spreading much more extensively (Fig. S8). Similarly, in the 2017/2018 A/H1N1pdm09 season, later onset for lineage 2 would have constrained it to the Pacific Northwest, whereas earlier onset would have facilitated substantially more expansive spread (Fig. S9).

### Mobility patterns coupled to inter-lineage competition explain differences in lineages’ spread

To further test the capacity of mobility-mediated inter-lineage competition to explain individual lineages’ spread, we performed in-depth investigations into the 2019/2020 B/Victoria season, which was characterized by anomalously high amounts of epidemic activity (*32*) and a highly spatially diverse lineage composition, with the largest lineages appearing to originate in or in the vicinity of California (lineage 1), Florida (lineage 2), Texas (lineage 3), Louisiana (lineage 4), Nevada (lineage 5), and Washington (lineage 6), respectively (Fig. 4). Some lineages spread to over half of all states (e.g. lineages 1 and 2 from Florida and California, respectively), whereas spread was more regional for others. We sought to establish if we could analogously reproduce the distribution of the lineages across states using epidemic simulations.

**Fig. 4:**
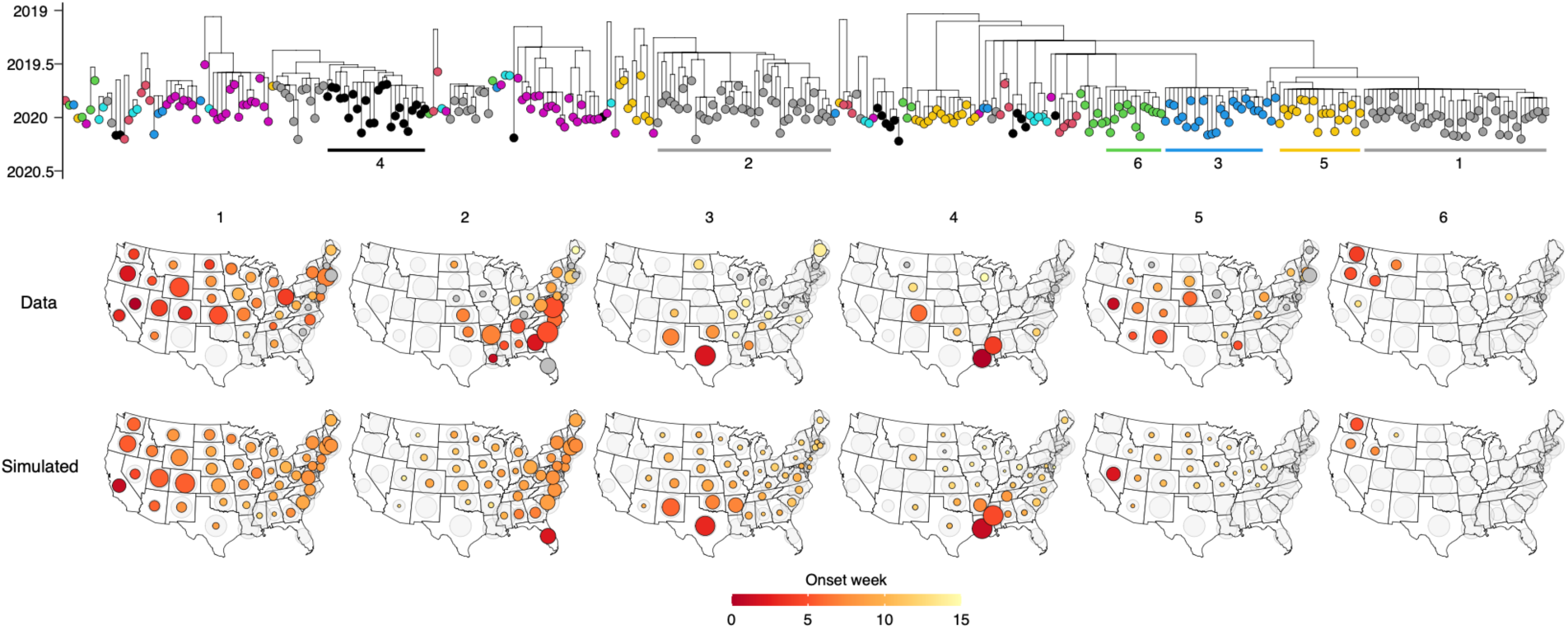
Mobility-induced competition drives individual lineage spread. Phylogeny represents the 2019/2020 B/Victoria season, with the six largest lineages labeled in order of size. Top row of maps represents the reconstructed spread and distribution of each of the six largest lineages. Bottom row of maps represents the simulated spread and distribution of the six lineages, initialized in the lineages’ respective onset state and onset week, simulated using a combination of air travel data and commuting data. Circle sizes are scaled as in Fig. 3.

Using a combination of commuting flows and air travel flows, the simulations reproduced the spread of individual lineages and their distribution across states (Fig. 4). Differences in mobility flows in combination with competition for susceptible individuals, linked to timing of lineage establishment, parsimoniously explain why the lineages emerging from California and Florida spread widely, whereas the lineages from Louisiana and Washington were more spatially constrained. Commuting flows in isolation provided a similarly strong fit, but underestimated spread from Nevada, suggesting that residual air travel flows not captured by commuting could play a role in viral dissemination (Fig. S10). Conversely, simulations using air travel deviated from the ground truth, particularly by underestimating short-range viral migration from Louisiana and Washington (Fig. S11).

The co-circulation of many lineages across distinct regions in this season illustrates how concurrent processes of epidemic establishment in different states, interacting with mobility, mediate nation-wide epidemic lineage composition and spatial structure. This season also highlights the heterogeneity of lineage establishment processes. For example, the lineage emerging from Florida likely emerged in in the spring of 2019 (posterior mean TMRCA May 5, 95% CrI March 14 –June 12), seemingly persisting throughout the 2019 summer in Florida. Hence, this lineage potentially provides a counterexample to the general trend that viruses do not persist between seasons (*24*). Conversely, the lineages from California, Nevada, and Texas spread widely following rapid establishment, despite much later emergence (e.g. lineage 1: posterior mean TMRCA August 29, 95% CrI July 3 – September 25).

### Rates of reconstructed viral migration correlate with commuting and not air travel

Using mechanistic metapopulation simulations, the above results suggest that commuting flows are the primary mobility drivers of influenza virus spread. However, these reconstructions could only be performed for the seasons with relatively low lineage diversity, as the large number of co-circulating lineages in some seasons rendered sample counts too low for individual lineages to yield a reliable ground truth for reconstructions of spread. To confirm that commuting is the predominant drivers of viral spread when incorporating all lineages across all seasons in the analysis, we investigated if the same mobility processes were reflected in the genetic relationships between viruses in the lineage phylogenetic trees themselves. To do so, we leveraged phylogeographic analyses to compute the relative role of each state as a donor or recipient state of viral migration events for each other state. Here, the relative viral jump contribution *x*→*y* represents the proportion of reconstructed migration events to and from state *y* that was accounted for by state *x*. Then, we correlated this metric of relative viral migration frequency with metrics of human mobility.

The states with the greatest role as the source or destination of viral migration events to/from a given state tended to be the states that were most strongly connected through commuting flows to that state (Spearman *ρ* = 0.63, *P* < 0.001) (Fig. 5A). A correlation between relative jump contribution and air travel contribution was also present (Spearman *ρ* = 0.32, *P* = <0.001), but this correlation was weaker, with often a relatively high pairwise viral migration frequency even for states poorly linked through air travel (Fig. 5B). These results provide orthogonal support for the dominant role of commuting flows in driving seasonal influenza virus spread. We note that because we use a maximally uninformative phylogeographic model for viral migration, the model likely overestimates rates of spatially uncorrelated spread, but our conclusions are robust to such biases (see Materials and Methods).

**Fig. 5:**
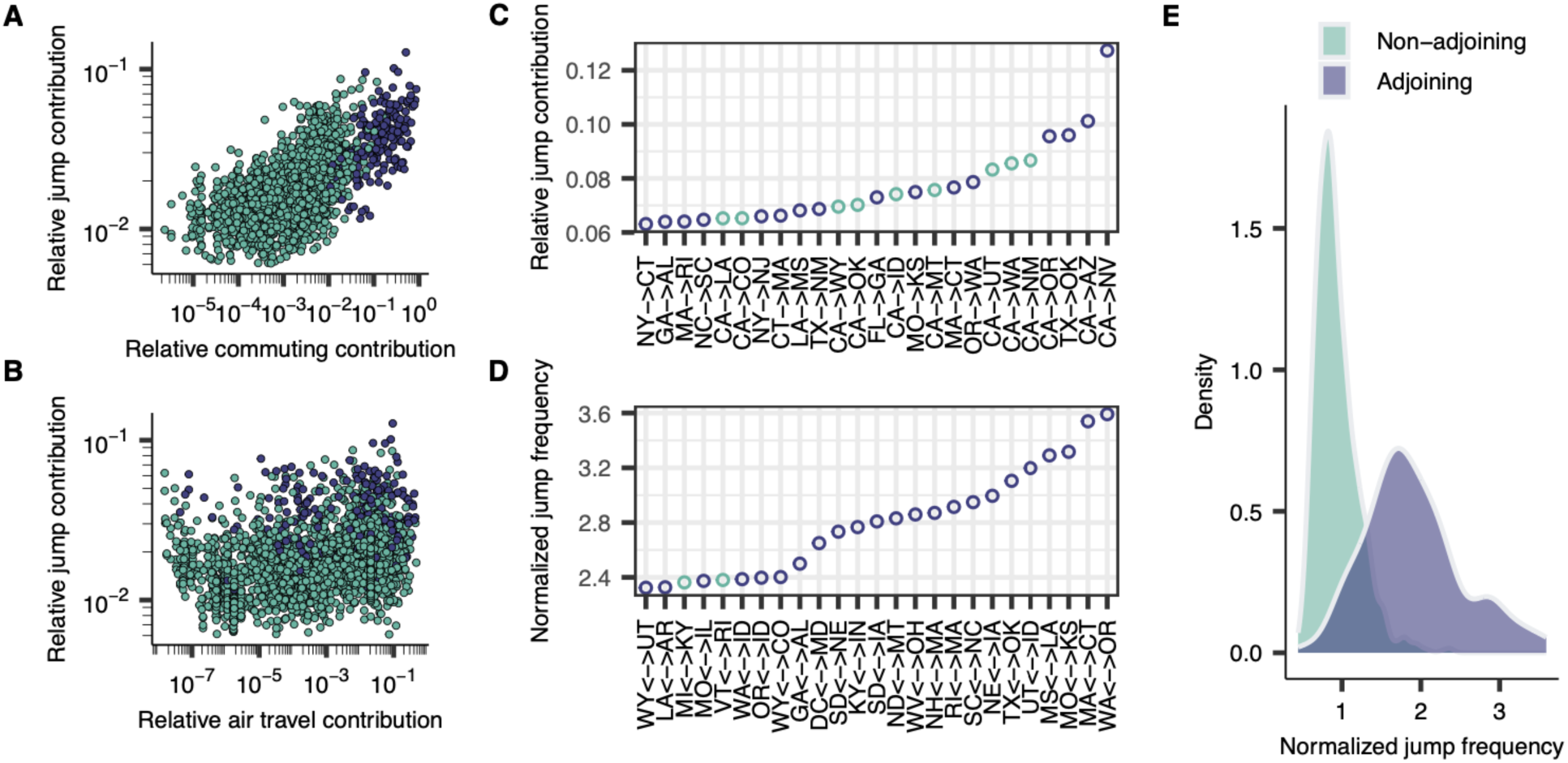
Phylogeographic analyses of mobility drivers. (A) Relationship between the relative contribution of each other state to a state’s inbound and outbound reconstructed viral migration events, and the other state’s relative role as a state’s commuting destination. (B) Analogous to (A), for air travel data. (C) Visualization of the 20 highest values of the relative jump contribution. (D) Visualization of the 20 highest values for the normalized pairwise jump frequency. (E) The distribution of normalized pairwise migration frequencies for pairs of adjoining and non-adjoining states.

The highest values of the relative jump contribution *x*→*y* were found when state *x* was highly populous and state *y* was in close geographical proximity to state *x*. For example, the highest values across all pairs were found for CA→NV, CA→AZ, TX→OK, CA→OR, and CA→NM (Fig. 5C). This is expected under classical gravity-like spread where, for any given state, the highest connectivity is expected to be to states that are in close geographic proximity and highly populous. However, this pattern could be confounded by higher sample counts for the most populous states, as higher sample counts for a deme in phylogeographic analyses will *a priori* be expected to lead to more reconstructed migration events even in the absence of any spatial signal in the data. As such, we also computed an alternative metric that accounts for this potential confounder. Here, the normalized pairwise jump frequency *x*↔*y* represents the proportion of migration events to/from state *y* that is accounted for by state *x*, normalized relative to the mean proportion of migration events that state *x* accounts for across all states. This quantity is symmetric, i.e. *x*↔*y* = *y*↔*x*. The highest values were for adjacent states that are strongly connected through commuting flows (highest: WA↔OR, MA↔CT, MO↔KS, MS↔LA, UT↔ID (Fig. 5D), indicating that when accounting for effects of population size and/or sampling, viral migration is strongly skewed toward short distances. Consistent with this, the normalized pairwise migration frequency was substantially greater for states that are adjacent than those that are non-adjacent (Fig. 5E). These results provide further evidence for the important role of short-distance spatial coupling in viral spread.

## Discussion

Our analyses at the transmission lineage level reveal the structure of seasonal influenza virus epidemics at a fine-grained resolution. Spread of individual lineages often occurred in a clear spatiotemporal hierarchy, and the competitive co-circulation of different lineages induced a strong spatial structure in seasonal influenza epidemics. The lineage structure of epidemics cannot reliably be identified from epidemiological data alone, but it is an essential component of seasonal influenza epidemiological dynamics. For example inter-state patterns of spatial coupling at the transmission lineage level as identified in this study differ substantially from patterns of similarity solely defined as correlation of influenza-like illness in approximately the same time period (*19*).

Furthermore, the previously identified strong spatial coupling between the more populous states from epidemic synchrony could be the result of concurrent processes of epidemic establishment resulting from distinct seeding events, rather than the result of hierarchical spread between different states (*5*, *26*). Lineage structure is also key to understanding seasonal influenza source-sink dynamics. Previous studies based on the ILI data have posited that the South represents the dominant source of influenza virus epidemics (*4*, *6*). While our analyses reveal the frequent early establishment and national success of lineages emerging in the South, this pattern was not consistent across seasons and the lineage complexity of epidemics means that source-sink dynamics are highly heterogeneous across seasons. Our inferences regarding source-sink dynamics also differ from those in studies that implicitly assume that a single lineage generated all epidemic activity in a given season (*33*).

Using a mechanistic epidemic model to reproduce lineage spread, we show that observed dynamics of lineage spread are mostly driven by commuting flows, which generate the network on which co-circulating lineages compete for disease-susceptible individuals. It is striking that we could reproduce lineage spread dynamics using mechanistic simulations when parameterizing mobility directly using commuter surveys. Commuting data has previously been suggested to drive influenza viral spread based on analyses of ILI data (*4*, *5*), but this has not been shown mechanistically or validated against phylogenetically supported instances of viral spread across (sub)types (*4*, *8*, *23*). While we found a clear dominance of commuting over air travel when considering these metrics in isolation, our results also suggested that air travel flows not captured in commuter surveys could play a role in viral dissemination.

The competitive dynamics of individual lineages exhibit the characteristics of gravity-like spread with localized, radial spread from less populous states, and strong long-range connections allowing rapid cross-country spread from highly populous states. The coupling induced by human mobility as reflected in the clear spatial hierarchies of lineage spread provides an explanation why including spatial coupling has been found to increase forecasting performance (*18*, *19*). The often highly repeatable dynamics of human mobility suggest a potential role for ensemble forecasts that integrate lineage-specific epidemic dynamics with patterns of human mobility to predict epidemic make-up. Upon local establishment of different lineages in different states, such simulations could be used to forecast which lineages are likely to predominate where, but given the stochastic dynamics of lineage establishment, forecasting efforts would also need to take into account uncertainties arising from the potentially rapid spread of lineages that have yet to establish. These forecasts could be especially valuable for public health planning if antigenically distinct viruses establish transmission chains in different states, such as observed in the 2018/2019 A/H3N2 season (*31*).

Early-season virologic surveillance data have been shown to given clues as to epidemic subtype composition (*20*), but the importance of short-term lineage establishment processes crucially suggests that the transmission chains corresponding to the earliest-sampled viruses will often not propagate into periods of peak epidemic activity, limiting the predictive utility of early-season genomic surveillance efforts. On the other hand, the strong correspondence between lineage establishment timing and lineage size, where the most dominant lineages are the ones that establish earliest, underscores the importance of high-resolution information on where substantial levels of seasonal influenza virus epidemic activity are occurring. Our study highlights the importance of nowcasting efforts to identify the locations of epidemic establishment which, when combined with high-resolution genomic surveillance in those areas, could be leveraged to generate more robust predictions of lineage spread (*19*).

Our analyses have a number of limitations. The procedure used to classify viruses into transmission lineages could introduce errors, but the strong spatial structures identified lend credence to the clustering method used. Furthermore, we could only perform our analyses at the state level owing to that being the level of spatial resolution in most virus metadata, and analyses at other spatial scales may yield different results regarding modes of virus spread (*4*, *34*). Our analyses are limited by the relatively low evolutionary rate and relatively limited sampling of influenza B viruses, complicating the accurate delineation into transmission lineages. Mobility flows underlying the spread of influenza B viruses are potentially different from those for influenza A viruses as a result of differences in the age distribution of infection (*2*). However, the identification of subtype-specific variation in dominant mobility flows was hampered by the substantial variation in epidemic size among subtypes and seasons, which renders potential differences in mobility flows difficult to disentangle from other sources of variation. Nevertheless, our mechanistic simulations were able to recapitulate observed patterns of spread using commuting data for influenza A and B viruses, suggesting similar mechanisms drive the spread of both.

We found that many of the most successful transmission lineages emerged very shortly before epidemic onset and established rapidly, sometimes sweeping to national dominance despite substantial competition from other contemporaneous transmission chains. The observed heterogeneity of transmission processes raises important questions regarding the predictability of early-season seasonal influenza epidemiological dynamics at multi-week time horizons, even in the presence of perfect data. The fact that seasonal influenza forecasts rarely outperform models based on historical baseline activity at timescales greater than a few weeks (*22*, *35*) is likely tied to these heterogeneities. An essential question remains what drives the timing and location of the highly explosive epidemic sparks that can lead to rapid lineage expansion. It is striking that in some seasons, the majority of peak-period circulation descended from a single ancestral virus that existed when relatively substantial circulation was already ongoing. Epidemic establishment processes are highly complex, likely influenced by many factors, including but not limited to immune susceptibility (*36–38*), climate (*11*, *39*, *40*), spatial organization (*11*), contact network structure (*10*), human behavior (*41*, *42*), inter-subtype competition (*37*, *43*), and international (*44*) and domestic travel acting in concert. High-resolution characterization of early-season epidemic dynamics at the transmission lineage level among diverse geographical localities is likely necessary to disentangle the contributions of these variables. Even if our results shed light on the potentially predictable underlying drivers of viral migration, an understanding of all the above factors will likely be necessary to probe the limits of seasonal influenza epidemic predictability.

## Materials & Methods

### Data

We downloaded all influenza A and B virus sequences corresponding to the A/H3N2 or A/H1N1pdm09 subtypes and B/Victoria and B/Yamagata influenza B lineages, collected from humans in the United States between July 1^st^ 2014 and July 1^st^ 2023 from the GISAID (*45*) EpiFlu database. Throughout, we use the term ‘subtype’ to refer to the influenza A subtypes and influenza B lineages individually, to avoid confusion between transmission lineages and influenza B lineages. We limited the dataset to viruses with sequences available for all eight gene segments. Furthermore, we retained only virus sequences with the US Centers for Disease Control as submitting laboratory, to minimize the impact of targeted sequencing investigations that are potentially not representative and could bias the data, particularly the branching structure of phylogenies. This led to a full dataset consisting of 30,508 viruses (A/H3N2: 14,235, A/H1N1pdm09: 8,155, B/Yamagata: 3,543, B/Victoria: 4,584). We downloaded weekly proportions reporting for influenza-like illness by state and season for the same time period from the CDC FluView website (https://www.cdc.gov/flu/weekly/fluviewinteractive.htm). From this website, we also downloaded weekly counts of positive influenza A and B tests in clinical laboratories, and weekly counts of positive tests by influenza A subtype and influenza B lineage in public health laboratories, by state and season. For the 2014/2015 season, positive tests for clinical laboratories were stratified by (sub)type/lineage. We similarly downloaded the number of positive tests in public health laboratories at the national level. We downloaded data on commuting flows for 2016-2020 from the US Census Bureau (https://www.census.gov/data/tables/2020/demo/metro-micro/commuting-flows-2020.html). This data is stratified by origin and destination county. We downloaded data on air travel fluxes between states, stratified by origin and destination airport for the year 2017 from the US Bureau of Transportation Statistics (https://www.transtats.bts.gov/DL_SelectFields.aspx?gnoyr_VQ=GED&QO_fu146_anzr=).

### Phylogenetic analyses

We aligned the sequences for each gene segment and subtype using MAFFT (*46*). We then clustered viruses, for each segment and subtype individually, into groups of highly related viruses using CD-HIT (*47*), with a clustering threshold of 99.5% nucleotide identity. Using a single representative virus for each cluster, we built a single tree for each subtype and segment individually using FastTree (*48*). We then fit a molecular clock to each tree using TempEst (*49*), and removed sequences belonging to CD-HIT- identified clusters for which the representative virus was classified as a molecular clock outlier from the dataset. This led to the removal of 40 viruses from the dataset. For each subtype, we then constructed a phylogenetic tree for each segment individually using all viruses for the entire period using IQTree (*50*) with a HKY (*51*) substitution model. We clustered the taxa in each of these trees by computing the largest groups of viruses where, for each taxon within a cluster, there was at least one other taxon in the group that saw a patristic distance to the former taxon that was smaller than a given distance threshold.

We defined this distance threshold as the expected number of mutations over a two-year period given the estimated molecular clock rate for that segment and subtype/lineage; we used a more relaxed three-year period for the MP and NS segments for additional lenience given their lower evolutionary rate. Using these cluster delineations, we assigned each taxon a segment-specific cluster identity. Using the cluster identities for each individual segment, we assigned each taxon a genome-wide cluster identity as the combination of individual segment identities. These identities were defined for each season individually, retaining the sequences from the 1^st^ of January of the preceding winter period up to the 1^st^ of July of the following year. We concatenated the sequences for all segments for viruses and constructed whole-genome phylogenetic trees for each of the genome-wide cluster identities individually. Concatenating gene segments runs the risk of introducing error due to potential reassortment events; we aimed to minimize this risk by clustering all taxa into groups of similar viruses using the procedure described above. We constructed phylogenies in IQTree (*50*) using a HKY (*51*) substitution model using a segment-proportional model (*52*). Given these maximum-likelihood phylogenies, we constructed temporally resolved trees using TreeTime (*53*) using a fixed clock rate estimated using TempEst.

### Transmission lineage identification

Using these time trees, we then sought to delineate the whole-genome phylogenetic trees into individual transmission lineages. We defined transmission lineages as groups of taxa on a phylogeny that plausibly descended from a common ancestor in the United States. Given the exponential nature of influenza epidemics, we identified groups of highly related viruses for which the tree structure follows the comb-like shape expected under an exponential growth population dynamic process, where most coalescent events happen close in time to the common ancestor. To do so, we used a modified version of Phydelity (*54*), a tool designed for the identification of transmission clusters on phylogenies. We imposed the constraint that each transmission lineage was required to exhibit the characteristic branching structure of exponential spread. Specifically, we required that for each transmission lineage, a certain proportion *p* of all coalescent events must occur within a particular period *t* of the putative lineage’s root, with *p* and *t* specified. Given the constraints, Phydelity aims to cluster as many taxa as possible given some constraints, formulating the problem as an integer linear programming (ILP) problem. Here, every internal node in the phylogeny is a potential transmission cluster, and the algorithm aims to cluster as many tips as possible, given the constraints.

If *t* is very high and *p* is very low the constraints imposed on the tree shape of a transmission lineage are relatively less stringent. As a result, sensitivity is high for purpose of clustering as many taxa as possible, but this might also result in erroneous clustering if genetically similar viruses were independently seeded into the United States and individually proliferated. On the other hand, a very stringent threshold (i.e. low *t* and high *p*) will lead to high specificity, but might also lead to erroneous discarding of true transmission lineages, as some true lineages will necessarily have a less comb-like structure, for example if they emerged early in the season, outside of typical periods of respiratory virus circulation, and spread at low levels before expanding when conditions were favorable for large-scale transmission. In the main text, we chose *p* = 0.10 and *t* = 1/12, i.e. 10% of coalescent events in a lineage must occur in the first month after its root. These values were chosen to balance sensitivity and specificity. We visualized the clustered trees using the *ggtree* (*55*) package, presenting all transmission lineages in a single tree. Because we clustered the trees into groups of similar viruses at the whole-genome level before the identification of transmission lineages, we did not reconstruct the ancestral relationships between all taxa. Hence, we only present the relationships between taxa if they belonged to the same whole-genome cluster identity. Differences in sampling among states could affect the delineations of viruses into clusters, principally by affect the branching structure within putative clusters. We found a strong log-linear relationship between a state’s population size and its sequencing rate relative to its population size (Pearson *r* = -0.73, *P* < 0.001), but some states had substantially greater sampling rates than would be expected under the identified relationship given their population size. To minimize effects of differences in sampling on cluster delineations, we subsampled the taxa in each state, for each season-subtype pair, such that no state had a number of sequences more than 0.5 log units greater than the regression-predicted number given its population size.

To reconstruct the spatiotemporal spread dynamics of individual lineages, we integrated the sampling date of each taxon with influenza-like illness and virological surveillance data. For each season and state individually, we computed the influenza type-specific disease signal by multiplying the proportion reporting influenza-like illness in each week by the proportion of tests positive for influenza A and B separately, yielding a measure of type-specific incidence. We then applied a 4253H, Twice smoother, implemented in the *sleekts* R package, to smooth the epidemic curves. To extract transmission lineage-specific epidemic curves, we fitted the sampling dates of taxa belonging to individual transmission lineages to the reconstructed type-specific epidemic curves. For each state and type (i.e. A or B) individually, we used a kernel density estimate given by a normal distribution centered around each taxon’s sampling date, with a two-week standard deviation. We retained taxa that were not assigned to any transmission lineage as a separate group. In each week, the relative incidence of a transmission lineage in that state was given by the proportion of all kernel density estimate contributions in that week corresponding to that lineage, multiplied by type-specific incidence.

Given each lineage’s reconstructed epidemic dynamics in each state, we computed two key state-level transmission lineage-specific summary statistics. 1) Lineage size, computed by dividing the number of taxa sampled in each state belonging to a particular by the total number of sequenced viruses in the state for that subtype, in the corresponding season, i.e. ranging from 0 to 1; and 2) lineage establishment timing, defined as the first week the lineage had accounted for at least 5% of total incidence in that season (if at all), using the mapping of sequence sampling date to incidence data described above. In some states, ILI and/or virologically confirmed data was absent for all or some seasons; in these cases, we only computed the lineage size, and not the onset week. Using these state-specific quantities, we computed nation-wide lineage size as the sum of state-specific relative sizes divided by the number of states; hence, each state is equal-weighted, irrespective of the state’s population size or sample count. We also computed the nation-wide time of lineage establishment as the first week of lineage establishment in any state. For each state, in each subtype-season pair, we computed the normalized Shannon entropy of the season’s lineage composition, which equates to 1 if each sampled virus corresponded to a different transmission lineage, and 0 if all sampled viruses belonged to the same lineage. In the regression analyses for the determinants of lineage size, we included only subtype-season pairs where the subtype accounted for >10% of a season’s total positive tests.

### Spread reconstruction

To characterize the similarity of lineage compositions across all pairs of states, we computed the median Bray-Curtis similarity for all pairs of states. We sampled 20 clustered viruses from each state for each season across all (sub)types (or retained all if fewer than 20 sequences were available) and computed the Bray-Curtis similarity of the transmission lineages corresponding to the sampled viruses using the *vegdist* command in the *vegan* (*56*) R package. We performed this procedure 50 times, retaining the mean value of each pair of states’ similarity across all replicates. All analyses were performed for the seasons from 2014/2015 to 2019/2020 and 2022/2023, omitting the 2021/2022 season due to its aberrant epidemic dynamics following the COVID-19 pandemic; this season saw substantial levels of circulation during the summer period, complicating the delineation of lineages into individual seasons. We retained only states with at least 10 sequences in all seasons, leaving a set of 42 states.

We performed hierarchical clustering on the similarity matrix across all seasons/subtypes using the *hclust* R function, using complete linkage clustering. We performed isometric multi-dimensional scaling using the *isoMDS* function in the *MASS* R package. To compute the correlation states between compositional similarity and centroid distance, we correlated the similarity matrix with states’ centroid distances using the *mantel* command in the *vegan* package. We performed these analyses at the individual season- subtype pair level in the same fashion, sampling 10 viruses from the set of clustered taxa sampled in that season for that subtype and computing the Bray-Curtis similarity as described above. Here, we retained only season-subtype pairs that saw at least 40 states with at least 10 clustered viruses.

### Source-sink phylogeographic inference

For the analyses of source-sink dynamics, we performed phylogeographic analyses in BEAST (*30*) for all transmission lineages that accounted for at least 0.5% of all sequenced viruses in a given season across all subtypes. We performed these analyses at the level of Health and Human Services (HHS) region, to allow for substantial spatial granularity while also having sufficient sequence counts per spatial unit. We used Thorney BEAST, implemented in BEAST (*30*) v2.3.31, to estimate a distribution of time-resolved phylogenies for each individual lineage, marginalizing over bifurcating topologies consistent with the potentially multifurcating input tree. We used divergence trees estimated in IQTREE, as explained above, as input trees, extracting the subtrees that corresponded to each transmission lineage. We furnished all transmission lineages with fewer than 50 taxa with an exponential growth coalescent prior, and a Skygrid (*57*) coalescent prior for all transmission lineages with at least 50 taxa. We estimated a single clock rate for each season–subtype pair. For each season–subtype pair, we ran a single MCMC chain for 500 million iterations, sampling lineage trees every 5 million states. We assessed convergence using Tracer (*58*), and generated a set of 90 posterior trees for each transmission lineage using TreeAnnotator (https://beast.community/treeannotator), removing the first 10% as burn-in.

We performed discrete trait phylogeographic inference (*59*) using the posterior lineage trees. We used a CTMC model for migration where we assumed equal rates of migration between all regions. We used this model as many lineages had relatively few sequences, prohibiting the reliable estimation of pairwise region-to-region migration rates.

Furthermore, these rates could not realistically be shared across transmission lineages as dynamics of migration vary substantially from lineage to lineage depending on the location of emergence and the landscape of lineage competition, and are likely highly temporally inhomogeneous, with dominance of the origin state early on but more spatially diffuse spread later. We ran these analyses for 100 million iterations, sampling every million, and removed the first 10% for each lineage as burn-in. We leveraged stochastic mapping (*60*) to identify migration events on the posterior phylogenies. Using these reconstructed migration events, we identified the likely origin of each lineage in each sample as the HHS region that was the source for most of the first 10 migration events in each lineage. We used this definition for the lineage origin rather than simply the reconstructed root region as we were primarily interested in the rapid expansion of lineages, and the root could be affected by the inclusion of unrelated singleton viruses in the analysis that did not contribute to lineage expansion. Reassuringly, we found that for most transmission lineages, lineage source posterior probabilities were generally focused on a small number of HHS regions.

Using the lineage origin posterior distributions, we then computed state-specific origin profiles, which represent the posterior proportion of sampled viruses in a focal state that belonged to lineages that were reconstructed to have originally expanded in each HHS region. Here, we aggregated across all subtypes and seasons, weighting each lineage according to the total proportion of circulation it accounted in the corresponding season in the focal state, across all subtypes. To investigate spatial structure in these source profiles, we correlated the Euclidean distance of these profiles between states with the centroid distance between the states using a Mantel test. Because we expected higher similarity between states in the same HHS region because they represent a single group in the phylogeographic reconstructions, we only performed this analysis for states that were not in the same HHS region. For any given state, we only included those seasons where that state had at least 10 sampled viruses when computing the source profiles, to prevent stochastic sampling effects from biasing results when sequence counts were low in a given season.

Phylogeographic reconstructions are prone to bias resulting from differences in sampling rates among the geographical groupings. To assess the sensitivity of our results with respect to these biases, we used two different sampling strategies. For our first sampling strategy, we used a sampling strategy where sequences from states that had a sequence count that was greater than expected from the regression line relating sequencing rate to population size were subsampled to the sequence count predicted from the regression line given its population size. This subsampling strategy was akin to the subsampling strategy used for the cluster delineations described above, but more stringent. Hence, the sample count for each HHS region was roughly proportional to the region’s population size. For the second sampling strategy, we ensured that the number of taxa included for each HHS region was approximately uniform, irrespective of the HHS region’s population size. For each season-subtype combination, we computed the sequence count as the 25^th^ quantile of the number of sequences in each HHS region in the population-proportional subsampling scheme used above. For regions with more sequences than this value, sequences were randomly subsampled. Because the results of the inferences are subject to variation due to the sampling strategy used, we mainly reported among-state differences for any given sampling strategy, and de-emphasized the absolute proportions estimated using the different models. For the same reason, we reported the likely origins of the largest lineages averaged across both sampling strategies. Nevertheless, the strong correlation between two sampling strategies suggests that sampling effects do not dominate the results.

To correlate mobility with rates of inter-state viral migration as reflected in the lineage phylogenies (Fig. 5), we performed Bayesian phylogeographic analyses analogous to the source-sink analyses above. We used the same procedure to perform phylogeographic reconstructions at the state level instead of the HHS region level, using the population-weighted subsampling strategy where sequences from states with higher sequencing rate than expected for their population were subsampled to the regression-predicted sequencing rate. We then reconstructed Markov jumps across all posterior trees for all transmission lineages. Then, for each pair of states *x* and *y*, we computed the relative jump contribution *x*→*y* as the proportion of migration events to and from state *y* that was accounted for by state *x*. We analogously computed the proportion of travelers from state *y* that had state *x* as destination for the air travel and commuting data and correlated these quantities with the relative jump contributions as estimated from the phylogenies. Here, we added a pseudocount for pairs of states with zero commuters or air travelers. We also computed the normalized relative jump frequency *x*↔*y*, which represents the proportion of migration events to/from state *y* that is accounted for by state *x*, normalized relative to the mean proportion of migration events that state *x* accounts for across all states. These values are highly symmetric (Pearson *r* = 0.997), and hence we symmetrized to subsume pairs of states. By comparing the jump frequency between states relative to the states’ mean, this metric is not prone to potential biases resulting from differences in sampling across states. However, a limitation of this metric is that only allows for ascertainment of the effect of distance and not of characteristics that are intrinsic to a single location, such as population size.

Because we used an equal-rates model for viral migration, the values of the relative jump contributions will likely overestimate rates of viral migration between spatially distant localities that are not well-connected through mobility. This is an inherent limitation of the model used. However, due to the complex migration dynamics that are likely highly time-inhomogeneous and differ substantially across lineages (as described above), migration patterns can likely not be captured by a single rate across all lineages and points in time, nor can time-inhomogeneous rates reliably be estimated or parameterized. These limitations also apply to alternative models such as a GLM formulation (*44*). Correlating reconstructed viral migration rates with metrics of mobility in *post hoc* analyses rather than including these metrics as covariates in the migration rate parameterizations affords certainty that the identified relationship between viral migration and human mobility is not a statistical artefact. The fact that we established a strong correlation between commuting rates and viral migration rates provides support for the use of the simplified model.

### Metapopulation model

With the aim of reproducing the observed spread of co-circulating lineages, particularly the lineages’ distribution among states, we used a mechanistic epidemic model that simulates the inter-state spread of co-circulating SIR-type pathogens with perfect cross-immunity that compete for disease-susceptible individuals. To limit the computational burden, we used a deterministic model that stratifies each epidemiological state into three further compartments for individuals remaining in their home state and for those visiting another state by commuting and air travel respectively. The model dynamics are then as follows:

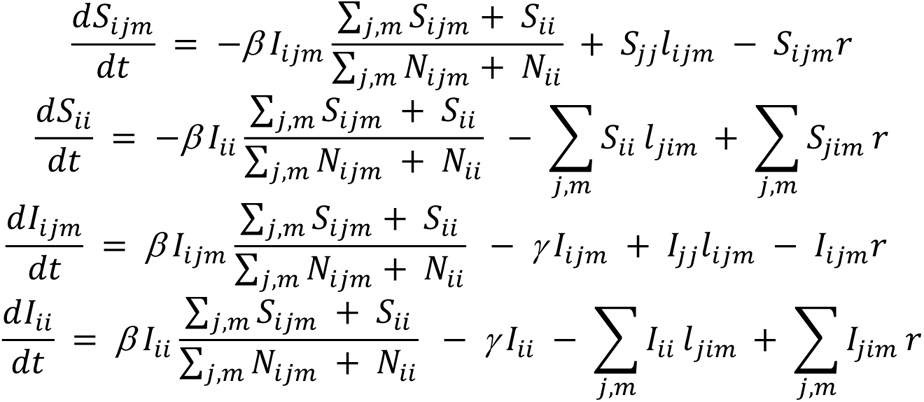

Here, *S_ijm_* and *I_ijm_* represent the number of susceptible and infected individuals, respectively, originating from state *j* that are currently in state *i* for mobility modality *m. m* can represent either commuting or air travel. Analogously, *S_jj_* and *I_jj_* represent the number of susceptible and infected individuals, respectively that are at home in state *j.* Matrix 𝑙 represent the outward travel rates for each mobility modality and *r* represents the return rate. We assume *r* = 1 day^-1^, *γ* = 0.25, and *R0* = 1.35. For state-to-state air travel rates, we computed the rate of air travel between states *x* and *y* as the total number of passengers in 2016 between airports located in state *x* and state *y*, and symmetrized the counts by computing the mean of the two counts. We then computed a daily rate from *x* to *y* by as the states’ symmetrized trip count divided by 365 and the population size of state *x*. We computed the number of commuters between each pair of states by aggregating across origin and destination counties in each state. We analogously symmetrized these counts (though they are highly symmetric, *r =* 0.9998) and computed the daily commuting rate *x*>*y* as the number of symmetrized commuters between the two states divided by the population size of state *x*. For the simulations that used a combination of air travel and commuting flows, the rate between each pair of states was defined as the maximum of the pairwise commuting and air travel rates, to account for the possibility that some of the commuting flows are accounted for by the air travel data. The model was implemented in C++, interfacing with R using Rcpp (*61*).

Using the metapopulation model, we investigated if we could recapitulate the distribution of lineages across the country, given the timing and location of each lineage’s onset, under the model of competition between lineages for susceptible individuals on the mobility network. For the set of lineages that were simulated, we simulated the epidemic progression forward in time, initializing each lineage in its reconstructed first week of establishment (see above), in its likely onset state. As the lineage’s onset week, we took the first week of establishment of the lineage in any state, rather than in the onset state, to account for situations where incidence data was absent for the likely onset state. However, we allowed each lineage’s onset week to vary to up to two weeks after or two weeks before its estimated data, to account for situations where the index state did not have incidence data available, and to account for error arising from the estimation of lineage-specific establishment timings with relatively noisy data. Lineages were initialized with an infected population of 1×10^-5^ times the index state’s population size. Analogous to the ground truth reconstructions, we computed the size of each lineage in the reconstructions as the proportion of infections across simulated lineages in a state that was attributable to a particular lineage. Similarly, we computed the week of establishment as the first week a lineage had caused >5% of total infections across simulated lineages in the full simulations. Because the simulations only included a limited set of lineages, we visualized the simulations by scaling the circles for each state such that the total size of the circles for the simulated lineages was proportional to the total proportion of sequences in each state that was accounted for by the simulated lineages.

## Data Availability

Sequence data is available from GISAID (accession numbers Table S1). All other data needed to evaluate the conclusions in the paper are present in the paper and/or the Supplementary Materials.

## Acknowledgments

We gratefully acknowledge the originating and submitting and laboratories that generated the sequence data that this study relies on.

## Author contributions

Conceptualization: SPJdJ, CAR

Methodology: SPJdJ, AC, AXH

Investigation: SPJdJ

Visualization: SPJdJ

Supervision: AXH, CAR

Writing—original draft: SPJdJ

Writing—review & editing: SPJdJ, AC, AXH, CAR

## Data and materials availability

Sequence data is available from GISAID (accession numbers Table S1). All other data needed to evaluate the conclusions in the paper are present in the paper and/or the Supplementary Materials. Code is available at http://www.github.com/AMC-LAEB/usa_flu.

## Supplementary Materials

Figs. S1 to S11

Table S1

## References

1. T. Bedford, S. Cobey, P. Beerli, M. Pascual, Global Migration Dynamics Underlie Evolution and Persistence of Human Influenza A (H3N2). PLOS Pathog. 6, e1000918- (2010).

2. T. Bedford, S. Riley, I. G. Barr, S. Broor, M. Chadha, N. J. Cox, R. S. Daniels, C. P. Gunasekaran, A. C. Hurt, A. Kelso, A. Klimov, N. S. Lewis, X. Li, J. W. McCauley, T. Odagiri, V. Potdar, A. Rambaut, Y. Shu, E. Skepner, D. J. Smith, M. A. Suchard, M. Tashiro, D. Wang, X. Xu, P. Lemey, C. A. Russell, Global circulation patterns of seasonal influenza viruses vary with antigenic drift. Nature 523, 217–220 (2015).

3. R. C. A, J. T. C, B. I. G, C. N. J, G. R. J, G. Vicky, G. I. D, H. A. W, H. A. J, H. A. C, de J. J. C, K. Anne, K. A. I, K. Tsutomu, K. Naomi, L. A. S, L. Y. P, M. Ana, O. Masatsugu, O. Takato, O. A. D. M. E, R. G. F, S. M. W, S. Eugene, S. Klaus, T. Masato, F. R. A. M, S. D. J, The Global Circulation of Seasonal Influenza A (H3N2) Viruses. Science (80-. ). 320, 340–346 (2008).

4. V. Charu, S. Zeger, J. Gog, O. N. Bjørnstad, S. Kissler, L. Simonsen, B. T. Grenfell, C. Viboud, Human mobility and the spatial transmission of influenza in the United States. PLOS Comput. Biol. 13, e1005382- (2017).

5. V. Cécile, B. O. N, S. D. L, S. Lone, M. M. A, G. B. T, Synchrony, Waves, and Spatial Hierarchies in the Spread of Influenza. Science (80-. ). 312, 447–451 (2006).

6. I. Chattopadhyay, E. Kiciman, J. W. Elliott, J. L. Shaman, A. Rzhetsky, Conjunction of factors triggering waves of seasonal influenza. Elife 7, e30756 (2018).

7. S. Pei, S. Kandula, W. Yang, J. Shaman, Forecasting the spatial transmission of influenza in the United States. Proc. Natl. Acad. Sci. 115, 2752 (2018).

8. B. A. Bozick, L. A. Real, The Role of Human Transportation Networks in Mediating the Genetic Structure of Seasonal Influenza in the United States. PLOS Pathog. 11, e1004898- (2015).

9. D. Balcan, V. Colizza, B. Gonçalves, H. Hu, J. J. Ramasco, A. Vespignani, Multiscale mobility networks and the spatial spreading of infectious diseases. Proc. Natl. Acad. Sci. 106, 21484 (2009).

10. S. Cauchemez, A.-J. Valleron, P.-Y. Boëlle, A. Flahault, N. M. Ferguson, Estimating the impact of school closure on influenza transmission from Sentinel data. Nature 452, 750– 754 (2008).

11. D. B. D, K. Stephen, G. J. R, V. Cecile, B. O. N, M. C. J. E, G. B. T, Urbanization and humidity shape the intensity of influenza epidemics in U.S. cities. Science (80-. ). 362, 75–79 (2018).

12. W. Yang, M. Lipsitch, J. Shaman, Inference of seasonal and pandemic influenza transmission dynamics. Proc. Natl. Acad. Sci. 112, 2723 (2015).

13. S. P. J. de Jong, Z. C. Felix Garza, J. C. Gibson, S. van Leeuwen, R. P. de Vries, G.-J. Boons, M. van Hoesel, K. de Haan, L. E. van Groeningen, K. D. Hulme, H. D. G. van Willigen, E. Wynberg, G. J. de Bree, A. Matser, M. Bakker, L. van der Hoek, M. Prins, N. A. Kootstra, D. Eggink, B. E. Nichols, A. X. Han, M. D. de Jong, C. A. Russell, Determinants of epidemic size and the impacts of lulls in seasonal influenza virus circulation. Nat. Commun. 15, 591 (2024).

14. N. G. Reich, L. C. Brooks, S. J. Fox, S. Kandula, C. J. McGowan, E. Moore, D. Osthus, E. L. Ray, A. Tushar, T. K. Yamana, M. Biggerstaff, M. A. Johansson, R. Rosenfeld, J. Shaman, A collaborative multiyear, multimodel assessment of seasonal influenza forecasting in the United States. Proc. Natl. Acad. Sci. 116, 3146 (2019).

15. J. Shaman, A. Karspeck, Forecasting seasonal outbreaks of influenza. Proc. Natl. Acad. Sci. 109, 20425 (2012).

16. M. Biggerstaff, M. Johansson, D. Alper, L. C. Brooks, P. Chakraborty, D. C. Farrow, S. Hyun, S. Kandula, C. McGowan, N. Ramakrishnan, R. Rosenfeld, J. Shaman, R. Tibshirani, R. J. Tibshirani, A. Vespignani, W. Yang, Q. Zhang, C. Reed, Results from the second year of a collaborative effort to forecast influenza seasons in the United States. Epidemics 24, 26–33 (2018).

17. M. Ben-Nun, P. Riley, J. Turtle, D. P. Bacon, S. Riley, Forecasting national and regional influenza-like illness for the USA. PLOS Comput. Biol. 15, e1007013- (2019).

18. A. E. L, N. A. T, V. Cecile, S. Mauricio, Toward the use of neural networks for influenza prediction at multiple spatial resolutions. Sci. Adv. 7, eabb1237 (2022).

19. F. S. Lu, M. W. Hattab, C. L. Clemente, M. Biggerstaff, M. Santillana, Improved state- level influenza nowcasting in the United States leveraging Internet-based data and network approaches. Nat. Commun. 10, 147 (2019).

20. E. Goldstein, S. Cobey, S. Takahashi, J. C. Miller, M. Lipsitch, Predicting the Epidemic Sizes of Influenza A/H1N1, A/H3N2, and B: A Statistical Method. PLOS Med. 8, e1001051- (2011).

21. D. Osthus, K. R. Moran, Multiscale influenza forecasting. Nat. Commun. 12, 2991 (2021).

22. C. Viboud, A. Vespignani, The future of influenza forecasts. Proc. Natl. Acad. Sci. 116, 2802–2804 (2019).

23. C. Viboud, M. I. Nelson, Y. Tan, E. C. Holmes, Contrasting the epidemiological and evolutionary dynamics of influenza spatial transmission. Philos. Trans. R. Soc. B Biol. Sci. 368, 20120199 (2013).

24. M. I. Nelson, L. Simonsen, C. Viboud, M. A. Miller, J. Taylor, K. S. George, S. B. Griesemer, E. Ghedin, N. A. Sengamalay, D. J. Spiro, I. Volkov, B. T. Grenfell, D. J. Lipman, J. K. Taubenberger, E. C. Holmes, Stochastic Processes Are Key Determinants of Short-Term Evolution in Influenza A Virus. PLOS Pathog. 2, e125- (2006).

25. M. I. Nelson, L. Edelman, D. J. Spiro, A. R. Boyne, J. Bera, R. Halpin, E. Ghedin, M. A. Miller, L. Simonsen, C. Viboud, E. C. Holmes, Molecular Epidemiology of A/H3N2 and A/H1N1 Influenza Virus during a Single Epidemic Season in the United States. PLOS Pathog. 4, e1000133 (2008).

26. P. Crépey, M. Barthélemy, Detecting Robust Patterns in the Spread of Epidemics: A Case Study of Influenza in the United States and France. Am. J. Epidemiol. 166, 1244–1251 (2007).

27. M. N. F, W. Cassia, F. C. D, R. Pavitra, L. Jover, M. L. H, P. Benjamin, R. Matthew, R. Erica, X. Hong, S. Lasata, A. Amin, R. V. M, L. N. A. P, H. Meei-Li, G. Romesh, M. Geoff, H. Brian, D. Philip, A. Amanda, B. Elisabeth, H. P. D, F. Kairsten, I. Misja, L. Kirsten, S. T. R, T. Melissa, W. C. R, B. Michael, E. J. A, F. Michael, L. B. R, R. M. J, T. Matthew, D. J. S, S. L. M, C. H. Y, S. Jay, J. K. R, L. Scott, G. A. L, N. D. A, B. Trevor, Viral genomes reveal patterns of the SARS-CoV-2 outbreak in Washington State. Sci. Transl. Med. 13, eabf0202 (2021).

28. D. Vijaykrishna, E. C. Holmes, U. Joseph, M. Fourment, Y. C. F. Su, R. Halpin, R. T. C. Lee, Y.-M. Deng, V. Gunalan, X. Lin, T. B. Stockwell, N. B. Fedorova, B. Zhou, N. Spirason, D. Kühnert, V. Bošková, T. Stadler, A.-M. Costa, D. E. Dwyer, Q. S. Huang, L. C. Jennings, W. Rawlinson, S. G. Sullivan, A. C. Hurt, S. Maurer-Stroh, D. E. Wentworth, G. J. D. Smith, I. G. Barr, The contrasting phylodynamics of human influenza B viruses. Elife 4, e05055 (2015).

29. B. I. Potter, R. Kondor, J. Hadfield, J. Huddleston, J. Barnes, T. Rowe, L. Guo, X. Xu, R. A. Neher, T. Bedford, D. E. Wentworth, Evolution and rapid spread of a reassortant A(H3N2) virus that predominated the 2017–2018 influenza season. Virus Evol. 5, vez046 (2019).

30. M. A. Suchard, P. Lemey, G. Baele, D. L. Ayres, A. J. Drummond, A. Rambaut, Bayesian phylogenetic and phylodynamic data integration using BEAST 1.10. Virus Evol. 4, vey016 (2018).

31. B. Flannery, R. J. G. Kondor, J. R. Chung, M. Gaglani, M. Reis, R. K. Zimmerman, M. P. Nowalk, M. L. Jackson, L. A. Jackson, A. S. Monto, E. T. Martin, E. A. Belongia, H. Q. McLean, S. S. Kim, L. Blanton, K. Kniss, A. P. Budd, L. Brammer, T. J. Stark, J. R. Barnes, D. E. Wentworth, A. M. Fry, M. Patel, Spread of Antigenically Drifted Influenza A(H3N2) Viruses and Vaccine Effectiveness in the United States During the 2018–2019 Season. J. Infect. Dis. 221, 8–15 (2020).

32. R. K. Borchering, C. E. Gunning, D. V Gokhale, K. B. Weedop, A. Saeidpour, T. S. Brett, P. Rohani, Anomalous influenza seasonality in the United States and the emergence of novel influenza B viruses. Proc. Natl. Acad. Sci. 118, e2012327118 (2021).

33. D. Magee, M. A. Suchard, M. Scotch, Bayesian phylogeography of influenza A/H3N2 for the 2014-15 season in the United States using three frameworks of ancestral state reconstruction. PLOS Comput. Biol. 13, e1005389 (2017).

34. S. Venkatramanan, A. Sadilek, A. Fadikar, C. L. Barrett, M. Biggerstaff, J. Chen, X. Dotiwalla, P. Eastham, B. Gipson, D. Higdon, O. Kucuktunc, A. Lieber, B. L. Lewis, Z. Reynolds, A. K. Vullikanti, L. Wang, M. Marathe, Forecasting influenza activity using machine-learned mobility map. Nat. Commun. 12, 726 (2021).

35. S. M. Mathis, A. E. Webber, T. M. León, E. L. Murray, M. Sun, L. A. White, L. C. Brooks, A. Green, A. J. Hu, R. Rosenfeld, D. Shemetov, R. J. Tibshirani, D. J. McDonald, S. Kandula, S. Pei, R. Yaari, T. K. Yamana, J. Shaman, P. Agarwal, S. Balusu, G. Gururajan, H. Kamarthi, B. A. Prakash, R. Raman, Z. Zhao, A. Rodríguez, A. Meiyappan, S. Omar, P. Baccam, H. L. Gurung, B. T. Suchoski, S. A. Stage, M. Ajelli, A. G. Kummer, M. Litvinova, P. C. Ventura, S. Wadsworth, J. Niemi, E. Carcelen, A. L. Hill, S. L. Loo, C. D. McKee, K. Sato, C. Smith, S. Truelove, S. Jung, J. C. Lemaitre, J. Lessler, T. McAndrew, W. Ye, N. Bosse, W. S. Hlavacek, Y. T. Lin, A. Mallela, G. C. Gibson, Y. Chen, S. M. Lamm, J. Lee, R. G. Posner, A. C. Perofsky, C. Viboud, L. Clemente, F. Lu, A. G. Meyer, M. Santillana, M. Chinazzi, J. T. Davis, K. Mu, A. Pastorey Piontti, A. Vespignani, X. Xiong, M. Ben-Nun, P. Riley, J. Turtle, C. Hulme-Lowe, S. Jessa, V. P. Nagraj, S. D. Turner, D. Williams, A. Basu, J. M. Drake, S. J. Fox, E. Suez, M. G. Cojocaru, E. W. Thommes, E. Y. Cramer, A. Gerding, A. Stark, E. L. Ray, N. G. Reich, L. Shandross, N. Wattanachit, Y. Wang, M. W. Zorn, M. Al Aawar, A. Srivastava, L. A. Meyers, A. Adiga, B. Hurt, G. Kaur, B. L. Lewis, M. Marathe, S. Venkatramanan, P. Butler, A. Farabow, N. Ramakrishnan, N. Muralidhar, C. Reed, M. Biggerstaff, R. K. Borchering, Title evaluation of FluSight influenza forecasting in the 2021–22 and 2022– 23 seasons with a new target laboratory-confirmed influenza hospitalizations. Nat. Commun. 15, 6289 (2024).

36. S. Gouma, K. Kim, M. E. Weirick, M. E. Gumina, A. Branche, D. J. Topham, E. T. Martin, A. S. Monto, S. Cobey, S. E. Hensley, Middle-aged individuals may be in a perpetual state of H3N2 influenza virus susceptibility. Nat. Commun. 11, 4566 (2020).

37. A. C. Perofsky, J. Huddleston, C. Hansen, J. R. Barnes, T. Rowe, X. Xu, R. Kondor, D. E. Wentworth, N. Lewis, L. Whittaker, B. Ermetal, R. Harvey, M. Galiano, R. S. Daniels, J. W. McCauley, S. Fujisaki, K. Nakamura, N. Kishida, S. Watanabe, H. Hasegawa, S. G. Sullivan, I. G. Barr, K. Subbarao, F. Krammer, T. Bedford, C. Viboud, Antigenic drift and subtype interference shape A(H3N2) epidemic dynamics in the United States. Cold Spring Harbor Laboratory (2023). 10.1101/2023.10.02.23296453.

38. T. Bedford, M. A. Suchard, P. Lemey, G. Dudas, V. Gregory, A. J. Hay, J. W. McCauley, C. A. Russell, D. J. Smith, A. Rambaut, Integrating influenza antigenic dynamics with molecular evolution. Elife 3, e01914 (2014).

39. J. Shaman, V. E. Pitzer, C. Viboud, B. T. Grenfell, M. Lipsitch, Absolute Humidity and the Seasonal Onset of Influenza in the Continental United States. PLOS Biol. 8, e1000316- (2010).

40. J. D. Tamerius, J. Shaman, W. J. Alonso, K. Bloom-Feshbach, C. K. Uejio, A. Comrie, C. Viboud, Environmental Predictors of Seasonal Influenza Epidemics across Temperate and Tropical Climates. PLOS Pathog. 9, e1003194- (2013).

41. Q. S. Huang, T. Wood, L. Jelley, T. Jennings, S. Jefferies, K. Daniells, A. Nesdale, T. Dowell, N. Turner, P. Campbell-Stokes, M. Balm, H. C. Dobinson, C. C. Grant, S. James, N. Aminisani, J. Ralston, W. Gunn, J. Bocacao, J. Danielewicz, T. Moncrieff, A. McNeill, L. Lopez, B. Waite, T. Kiedrzynski, H. Schrader, R. Gray, K. Cook, D. Currin, C. Engelbrecht, W. Tapurau, L. Emmerton, M. Martin, M. G. Baker, S. Taylor, A. Trenholme, C. Wong, S. Lawrence, C. McArthur, A. Stanley, S. Roberts, F. Rahnama, J. Bennett, C. Mansell, M. Dilcher, A. Werno, J. Grant, A. van der Linden, B. Youngblood, P. G. Thomas, R. J. Webby, Npi. Consortium, Impact of the COVID-19 nonpharmaceutical interventions on influenza and other respiratory viral infections in New Zealand. Nat. Commun. 12, 1001 (2021).

42. A. C. Perofsky, C. L. Hansen, R. Burstein, S. Boyle, R. Prentice, C. Marshall, D. Reinhart, B. Capodanno, M. Truong, K. Schwabe-Fry, K. Kuchta, B. Pfau, Z. Acker, J. Lee, T. R. Sibley, E. McDermot, L. Rodriguez-Salas, J. Stone, L. Gamboa, P. D. Han, A. Adler, A. Waghmare, M. L. Jackson, M. Famulare, J. Shendure, T. Bedford, H. Y. Chu, J. A. Englund, L. M. Starita, C. Viboud, Impacts of human mobility on the citywide transmission dynamics of 18 respiratory viruses in pre- and post-COVID-19 pandemic years. Nat. Commun. 15, 4164 (2024).

43. E. K. S. Lam, D. H. Morris, A. C. Hurt, I. G. Barr, C. A. Russell, The impact of climate and antigenic evolution on seasonal influenza virus epidemics in Australia. Nat. Commun. 11, 2741 (2020).

44. P. Lemey, A. Rambaut, T. Bedford, N. Faria, F. Bielejec, G. Baele, C. A. Russell, D. J. Smith, O. G. Pybus, D. Brockmann, M. A. Suchard, Unifying Viral Genetics and Human Transportation Data to Predict the Global Transmission Dynamics of Human Influenza H3N2. PLOS Pathog. 10, e1003932- (2014).

45. Y. Shu, J. McCauley, GISAID: Global initiative on sharing all influenza data - from vision to reality. (2017). 10.2807/1560-7917.ES.2017.22.13.30494.

46. K. Katoh, D. M. Standley, MAFFT Multiple Sequence Alignment Software Version 7: Improvements in Performance and Usability. Mol. Biol. Evol. 30, 772–780 (2013).

47. W. Li, A. Godzik, Cd-hit: a fast program for clustering and comparing large sets of protein or nucleotide sequences. Bioinformatics 22, 1658–1659 (2006).

48. M. N. Price, P. S. Dehal, A. P. Arkin, FastTree 2 – Approximately Maximum-Likelihood Trees for Large Alignments. PLoS One 5, e9490 (2010).

49. A. Rambaut, T. T. Lam, L. Max Carvalho, O. G. Pybus, Exploring the temporal structure of heterochronous sequences using TempEst (formerly Path-O-Gen). Virus Evol. 2, vew007 (2016).

50. B. Q. Minh, H. A. Schmidt, O. Chernomor, D. Schrempf, M. D. Woodhams, A. von Haeseler, R. Lanfear, IQ-TREE 2: New Models and Efficient Methods for Phylogenetic Inference in the Genomic Era. Mol. Biol. Evol. 37, 1530–1534 (2020).

51. M. Hasegawa, H. Kishino, T. Yano, Dating of the human-ape splitting by a molecular clock of mitochondrial DNA. J. Mol. Evol. 22, 160–174 (1985).

52. D. A. Duchêne, K. J. Tong, C. S. P. Foster, S. Duchêne, R. Lanfear, S. Y. W. Ho, Linking Branch Lengths across Sets of Loci Provides the Highest Statistical Support for Phylogenetic Inference. Mol. Biol. Evol. 37, 1202–1210 (2020).

53. P. Sagulenko, V. Puller, R. A. Neher, TreeTime: Maximum-likelihood phylodynamic analysis. Virus Evol. 4, vex042 (2018).

54. A. X. Han, E. Parker, S. Maurer-Stroh, C. A. Russell, Inferring putative transmission clusters with Phydelity. Virus Evol. 5, vez039 (2019).

55. G. Yu, D. K. Smith, H. Zhu, Y. Guan, T. T.-Y. Lam, ggtree: an r package for visualization and annotation of phylogenetic trees with their covariates and other associated data. Methods Ecol. Evol. 8, 28–36 (2017).

56. P. Dixon, VEGAN, a package of R functions for community ecology. J. Veg. Sci. 14, 927–930 (2003).

57. V. Hill, G. Baele, Bayesian Estimation of Past Population Dynamics in BEAST 1.10 Using the Skygrid Coalescent Model. Mol. Biol. Evol. 36, 2620–2628 (2019).

58. A. Rambaut, A. J. Drummond, D. Xie, G. Baele, M. A. Suchard, Posterior Summarization in Bayesian Phylogenetics Using Tracer 1.7. Syst. Biol. 67, 901–904 (2018).

59. P. Lemey, A. Rambaut, A. J. Drummond, M. A. Suchard, Bayesian Phylogeography Finds Its Roots. PLOS Comput. Biol. 5, e1000520 (2009).

60. M. A. R. Ferreira, M. A. Suchard, Bayesian analysis of elapsed times in continuous-time Markov chains. Can. J. Stat. 36, 355–368 (2008).

61. D. Eddelbuettel, R. Francois, Rcpp: Seamless R and C++ Integration. J. Stat. Softw. 40, 1–18 (2011).

