## Supplementary Materials for "Commuting-driven competition between transmission chains shapes seasonal influenza virus epidemics in the United States"

Science Advances

AAAS

Simon P.J. de Jong *et al.*

**This PDF file includes:**

Figs. S1 to S11

**Other Supplementary Materials for this manuscript include the following:**

Table S1

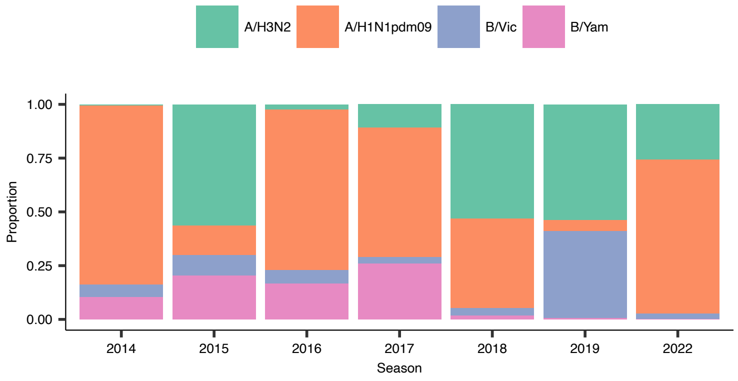


Fig. S1. Nation-wide epidemic compositions for the seasons included in the analysis, as computed from virological surveillance data.

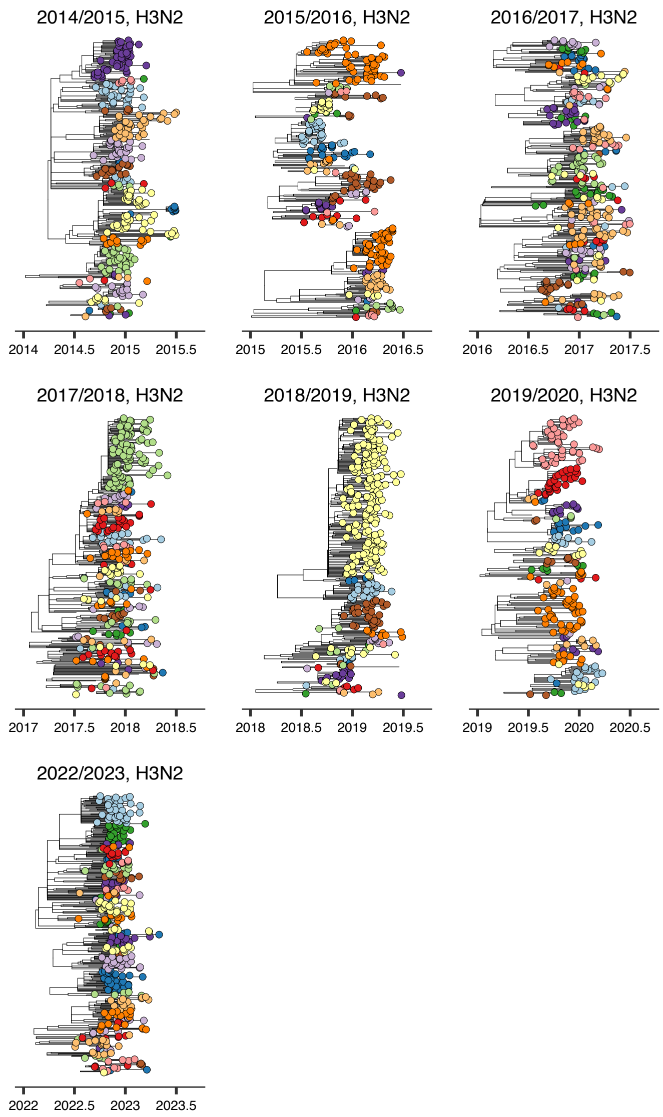


Fig S2. Phylogenies for the A/H3N2 subtype, clustered by transmission lineage.

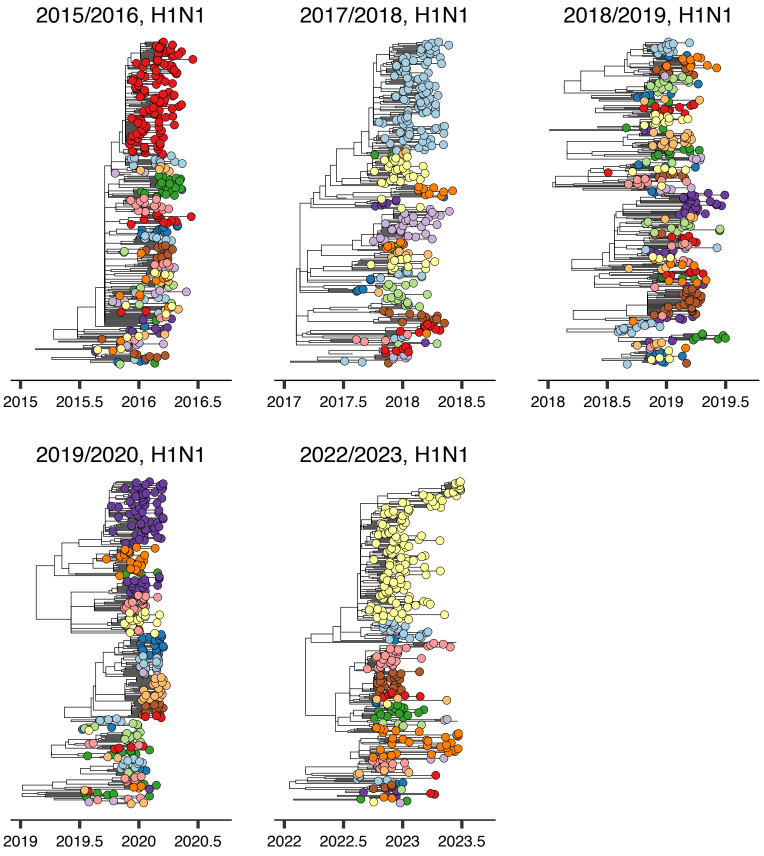


Fig S3. Phylogenies for the A/H1N1pdm09 subtype, clustered by transmission lineage.

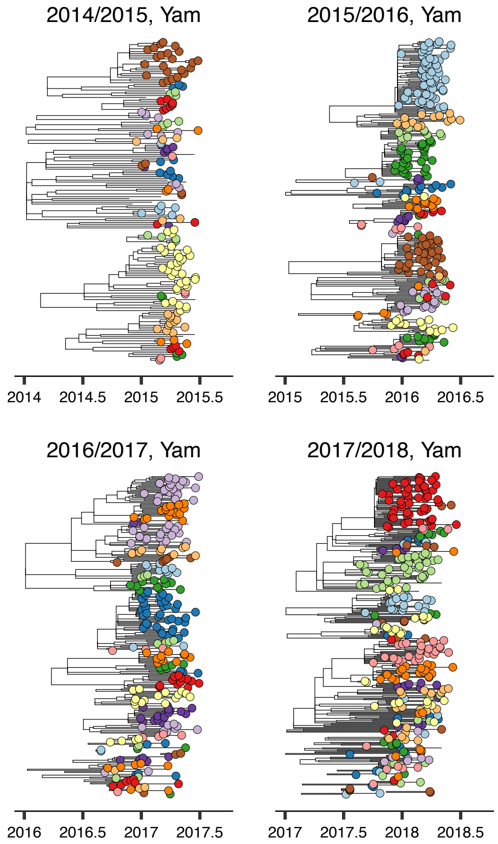


Fig S4. Phylogenies for the B/Yamagata subtype, clustered by transmission lineage.

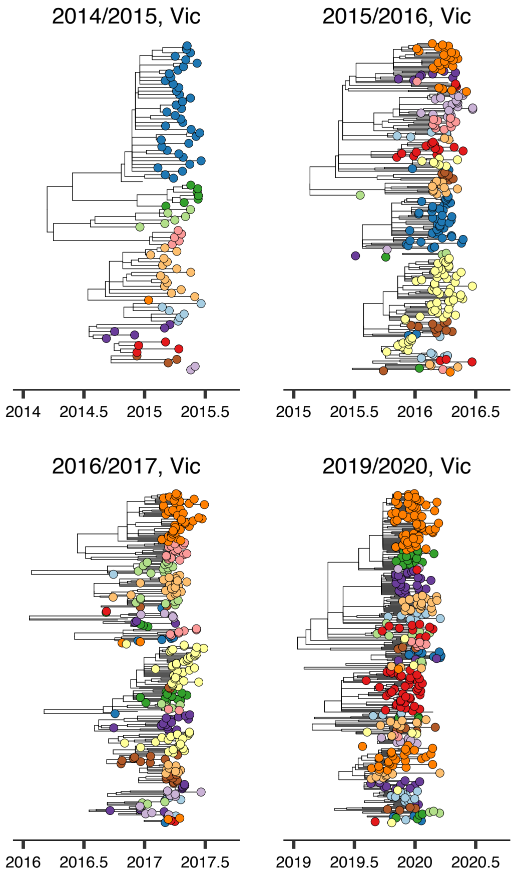


Fig S5. Phylogenies for the B/Victoria subtype, clustered by transmission lineage.

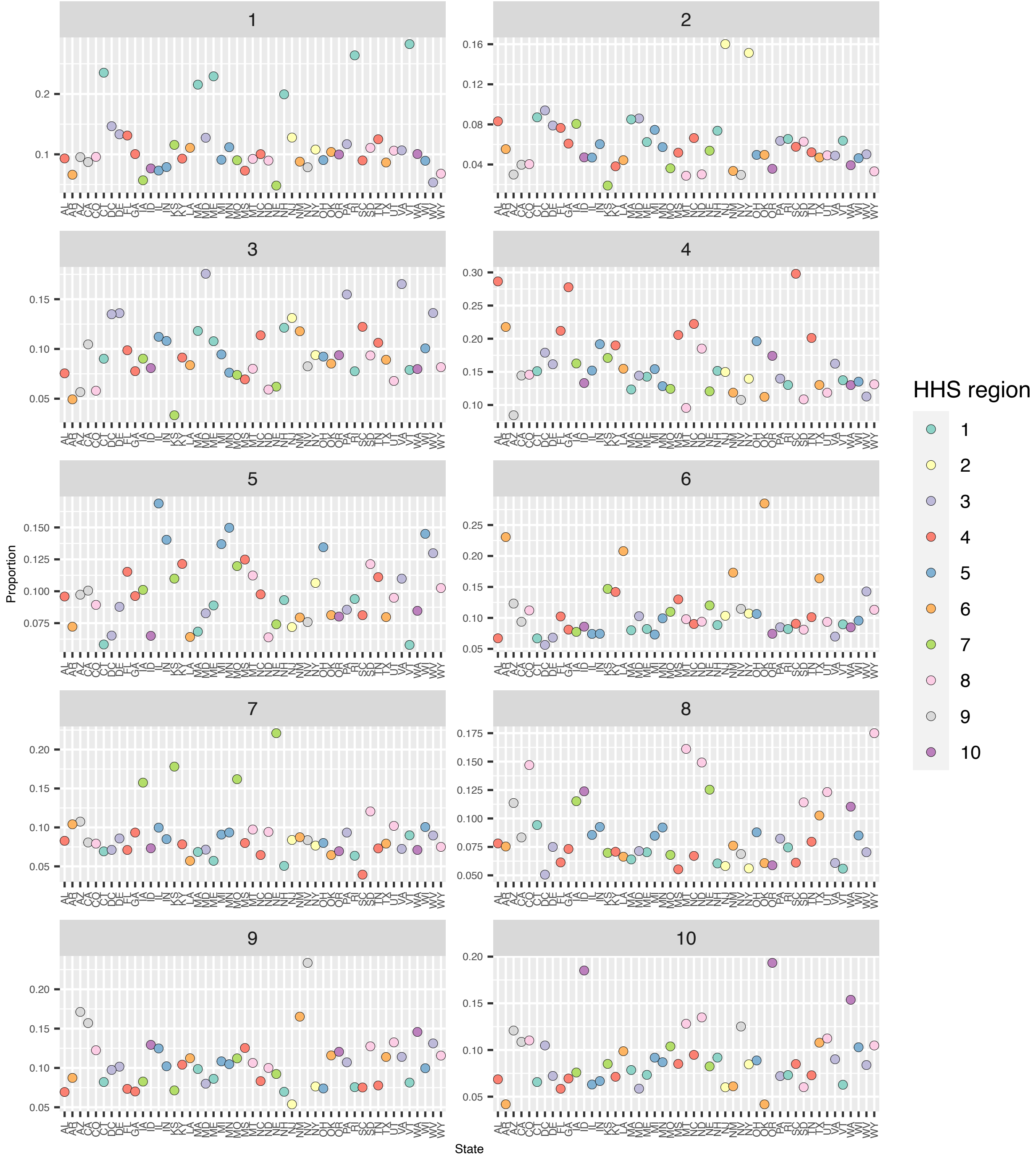


Fig S6. The proportion (*y*-axis) of sequences in each state (*x*-axis) that can be attributed to lineages that originally expanded from each HHS region. Each subpanel corresponds to a source HHS region. Results are for the subsampling strategy with uniform sampling across HHS regions.

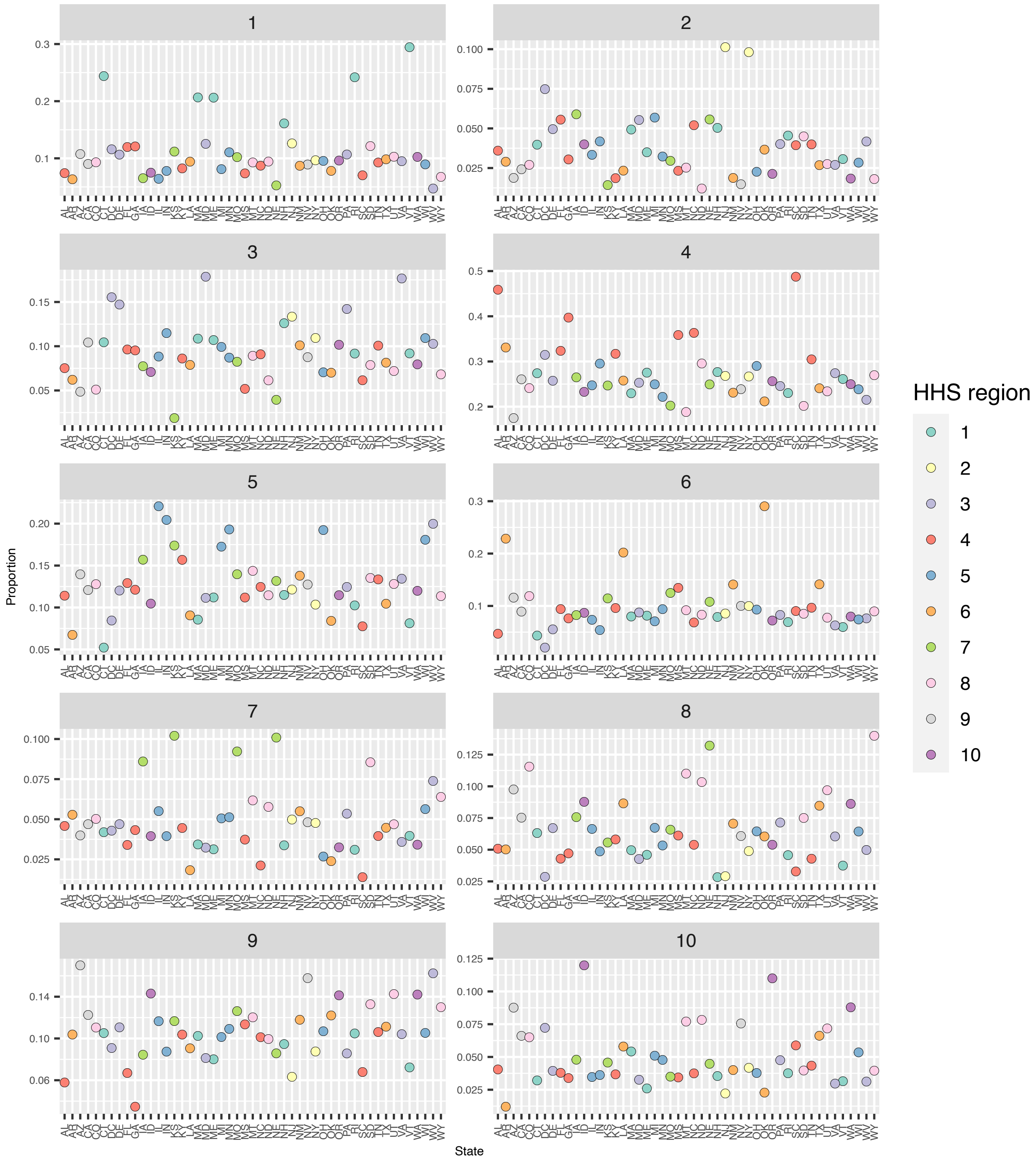


Fig S7. The proportion ($y$-axis) of sequences in each state ($x$-axis) that can be attributed to lineages that originally expanded from each HHS region. Each subpanel corresponds to a source HHS region. Results are for the subsampling strategy with population size-based sampling across HHS regions.

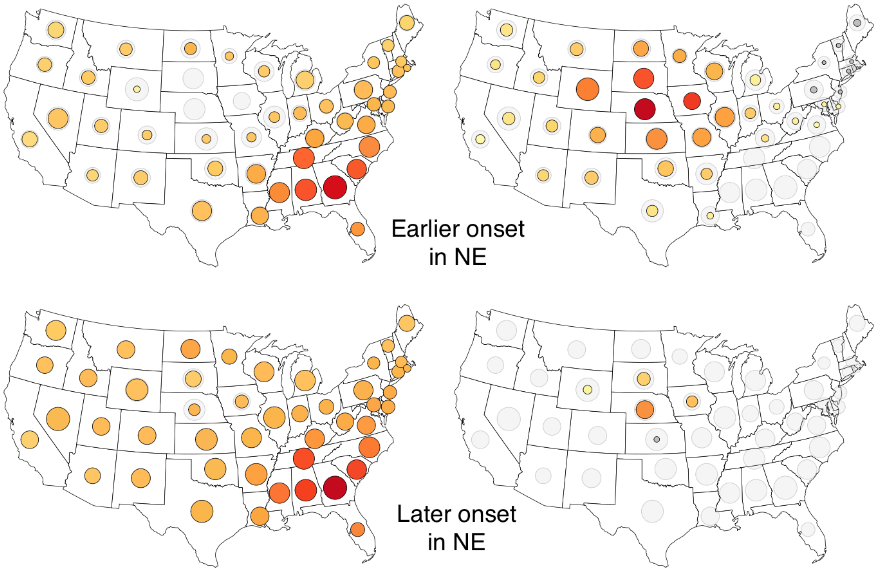


Fig S8: The simulated spread of the two largest lineages in the 2018/2019 A/H3N2 season, had lineage 2 established four weeks sooner (top) or four weeks later (bottom). Light grey circles represent the total proportion of sequences in that state that are accounted for by the lineages that were simulated, to account for the fact that simulations only incorporated a subset of all lineages; circles for the simulated lineages have their size scaled such that the sum of simulated lineages' sizes for each state is proportional to the proportion of sequences accounted for by the simulated lineages in that state (i.e., the light grey area). Dark grey fill corresponds to absence of an establishment week (for top row, potentially due to missing data), or establishment after the 15th week.

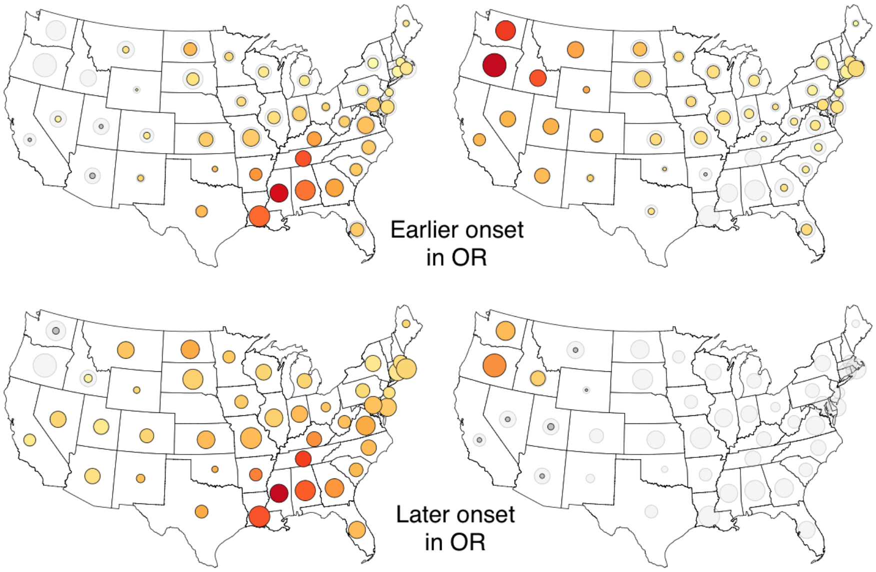


Fig S9: The simulated spread of the two largest lineages in the 2017/2018 A/H1N1pdm09 season, had lineage 2 established four weeks sooner (top) or four weeks later (bottom). Light grey circles represent the total proportion of sequences in that state that are accounted for by the lineages that were simulated, to account for the fact that simulations only incorporated a subset of all lineages; circles for the simulated lineages have their size scaled such that the sum of simulated lineages' sizes for each state is proportional to the proportion of sequences accounted for by the simulated lineages in that state (i.e., the light grey area). Dark grey fill corresponds to absence of an establishment week (for top row, potentially due to missing data), or establishment after the 15th week.

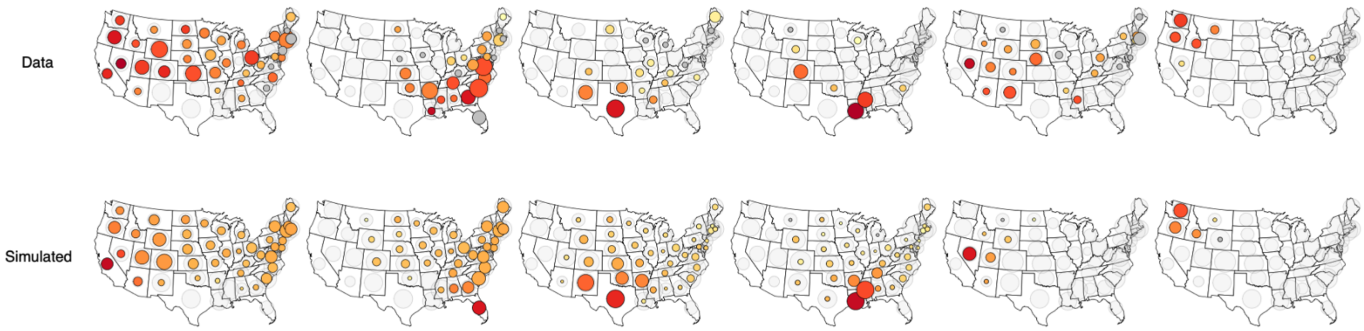


Fig S10: Top row of maps represents the reconstructed spread and distribution of each of the six largest lineages in the 2019/2020 B/Victoria season. Bottom row of maps represents the simulated spread and distribution of the six lineages, initialized in the lineages' respective onset state and onset week, simulated using commuting data. Circle sizes are scaled as in Fig. 3.

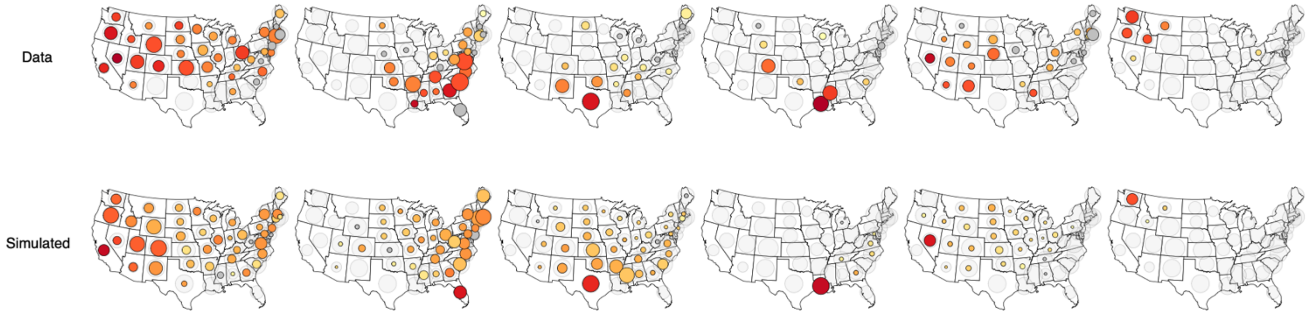


Fig S11: Top row of maps represents the reconstructed spread and distribution of each of the six largest lineages in the 2019/2020 B/Victoria season. Bottom row of maps represents the simulated spread and distribution of the six lineages, initialized in the lineages' respective onset state and onset week, simulated using air travel data. Circle sizes are scaled as in Fig. 3.
